# Analysis of Glomerular Transcriptomes from Nephrotic Patients Suggest *APOL1* Risk Variants Impact Parietal Epithelial Cells

**DOI:** 10.1101/2024.11.05.24316766

**Authors:** Agustin Gonzalez-Vicente, Dana C. Crawford, William S. Bush, Zhenzhen Wu, Leslie A. Bruggeman, Viji Nair, Felix Eichinger, Oliver Wessely, Matthias Kretzler, John F. O’Toole, John R. Sedor, Kidney Precision Medicine Project, Nephrotic Syndrome Study Network

## Abstract

The disproportionate risk for idiopathic proteinuric podocytopathies in Black people is explained, in part, by the presence of two risk alleles (G1 or G2) in the *APOL1* gene. The pathogenic mechanisms responsible for this genetic association remain incompletely understood. We analyzed glomerular RNASeq transcriptomes from patients with idiopathic nephrotic syndrome of which 72 had inferred African ancestry (AA) and 152 did not (noAA). Using gene coexpression networks we found a significant association between *APOL1* risk allele number and the coexpression metamodule 2 (MM2), even after adjustment for eGFR and proteinuria at biopsy. Unadjusted Kaplan-Meier curves showed that unlike noAA, AA with the highest tertile of MM2 gene activation scores were less likely to achieve complete remission (p≤0.014). Characteristic direction (ChDir) identified a signature of 1481 genes, which separated patients with *APOL1* risk alleles from those homozygous for reference *APOL1*. Only in AA, the tertile with the highest activation scores of these 1481 genes was less likely to achieve complete remission (p≤0.022) and showed a trend to faster progression to the composite event of kidney failure or loss of 40% eGFR (p≤0.099). The MM2 and ChDir genes significantly overlapped and were both enriched for Epithelial Mesenchymal Transition and inflammation terms. Finally, MM2 significantly overlapped with a parietal epithelial cell (PEC)-identity gene signature but not with a podocyte identity signature. Podocytes expressing variant APOL1s may generate inflammatory signals that activate PECs by paracrine mechanisms contributing to *APOL1* nephropathy.

## INTRODUCTION

Progressive chronic kidney diseases (CKD) are more common among Black people than other populations. The excess risk for non-diabetic CKD is associated with 2 *APOL1* variants (G1 and G2), unique to some African ancestral populations, under a recessive model of inheritance. The incomplete penetrance of the *APOL1* risk genotypes is consistent with disease modifiers, mostly undefined, but including recently discovered genetic modifiers [1–3] and high interferon states that drive APOL1 expression [4]. Induced kidney risk APOL1s, but not reference *APOL1* (G0), insert into the plasma membrane and form non-selective cation channels. The resulting sodium and potassium fluxes or the intracellular calcium increase result in activation of incompletely characterized cytotoxic pathways [5–9]. Animal models to study *APOL1* kidney disease mechanisms are limited to transgenesis, since only humans and some non-human primates carry the *APOL1* gene. Mice expressing *APOL1* risk variants, but not G0 transgenes, by either podocyte-specific tetracycline inducible expression systems, human *APOL1*-containing fosmids, or bacterial artificial chromosomes, develop kidney injury [10–12]. These findings establish a causal link between the expression of *APOL1* risk variants and kidney disease, but do not clearly define the mechanisms underlying the kidney injury [10–12]. G1, but not G0, kidney organoids, treated with interferon-γ to induce *APOL1* expression and exposed to tunicamycin to simulate a stress, demonstrated epithelial dedifferentiation but remained viable [13]. Most animal and *in vitro* studies demonstrating cytotoxic mechanisms fail to model a triggering stress; instead, *APOL1* is ectopically expressed which might short-circuit endogenous regulatory pathways that mitigate *APOL1*-dependent cytotoxicity. To explore *APOL1* pathobiology in humans, we analyzed the glomerular transcriptomes from the Nephrotic Syndrome Study Network (NEPTUNE) kidney biopsy cohort, which includes participants with focal segmental glomerulosclerosis (FSGS), steroid resistant nephrotic syndrome and membranous nephropathy. Since *APOL1* risk genotypes are strongly associated with idiopathic proteinuric podocytopathies, we hypothesized that “individuals with *APOL1* risk-alleles have a glomerular transcriptional signature, which could identify candidate disease mechanisms”. A previous study of *APOL1* using the NEPTUNE glomerular transcriptomes was limited to 30 self-identified or genotype-predicted Black subjects with a histologic diagnosis of FSGS [14]. Clinical glomerular histology poorly aligns with molecular mechanisms [15]. In addition, *APOL1* risk genotypes associate with steroid-resistant nephrotic syndrome and idiopathic membranous nephropathy [16–19]. Given this, we included all NEPTUNE subjects with glomerular transcriptomes and *APOL1* genotypes validated by DNA sequencing, regardless of the clinical histopathological diagnosis.

## RESULTS

We used a systems biology approach to study the pathobiology of *APOL1* kidney risk variants in a cohort of patients with nephrotic syndrome. An overview of the methodology is presented in **Supplemental Figure 1**.

### Clinical characteristics of the cohort (**Table 1**)

NEPTUNE participants with glomerular transcriptomes and validated *APOL1* genotypes (n=224) were included in these analyses; 72 participants had inferred (see Methods) African Ancestry (AA) and 152 individuals reported other ancestries (noAA). AA participants had lower eGFRs (76 ± 37 vs 95 ± 43 ml/min/1.73m^2^; p≤0.001) and were significantly more likely to be hypertensive at time of biopsy (p≤0.01). Proteinuria in AA was marginally lower compared to noAA participants (p≤0.054). In AA, kidney biopsy diagnosis was more frequently FSGS and less commonly minimal change disease (MCD) or membranous nephropathy (MN). The distributions of *APOL1* alleles stratified by the kidney biopsy nephrotic syndrome diagnosis are shown in **Supplemental Table 1**. Similar numbers of AA and noAA participants were treated with immunosuppressants, or drugs that inhibited the renin angiotensin aldosterone system.

**Table 1:** Participant characteristics stratified by inferred African ancestry (AA). Inferred AA was defined as either having an *APOL1* genotype different from G0/G0 or a self-reported race of “Black/African American” (NEPTUNE classification). Individuals homozygous for the *APOL1* G0 allele, who did not self-identified as “Black/African American”, were designated as not having inferred African ancestry (noAA). Categorical variables are expressed as percentages, with p-values calculated using Fisher’s exact test. Age, eGFR and log(1 + UPCR [urinary protein creatinine ratio]) (UPCR_lg) are expressed as mean ± SD. The Shapiro-Wilk test showed age (p≤1×10^−9^), eGFR (p≤2×10^−6^) and UPCR_lg (p≤2×10^−16^) significantly deviated from a normal distribution and the Wilcoxon rank sum test determined if differences between AA and noAA were significant. Subscripts (in parenthesis) are the numbers of individuals (n) in each group, which differs slightly between phenotypes due to missing data points.

|  | **AA (72)** | **noAA (152)** | **p value** |
| --- | --- | --- | --- |
| Sex male (%) | 64%<sub>(46)</sub> | 63%<sub>(95)</sub> | *0.883* |
| Pediatric (%) | 42%<sub>(30)</sub> | 36%<sub>(54)</sub> | *0.380* |
| Hypertension (%) | 61%<sub>(44)</sub> | 42%<sub>(64)</sub> | ***0.010*** |
| Diabetes (%) | 4%<sub>(3)</sub> | 7%<sub>(10)</sub> | *0.057* |
| Immunosuppression (%) | 36%<sub>(26)</sub> | 44%<sub>(67)</sub> | *0.310* |
| ACE/ARB (%) | 32%<sub>(23)</sub> | 33%<sub>(50)</sub> | *1.000* |
| Age at biopsy (years) | 31 ± 21<sub>(72)</sub> | 34 ± 23<sub>(152)</sub> | *0.600* |
| eGFR (mL/min/1.73m<sup>2</sup>) | 76 ± 37<sub>(70)</sub> | 95 ± 43<sub>(140)</sub> | ***0.001*** |
| UPCR<sub>lg</sub> | 0.59 ± 0.36<sub>(71)</sub> | 0.69 ± 0.37<sub>(146)</sub> | *0.054* |
| **Cohort (%)** |  |  |  |
| FSGS | 49%<sub>(35)</sub> | 34%<sub>(52)</sub> | - |
| MCD | 33%<sub>(24)</sub> | 38%<sub>(58)</sub> | - |
| MN | 18%<sub>(13)</sub> | 28%<sub>(42)</sub> | - |
| **Race (%)** |  |  |  |
| Black/African American | 85%<sub>(61)</sub> | 0 | - |
| Multi-Racial | 10%<sub>(7)</sub> | 3%<sub>(5)</sub> | - |
| Unknown | 2.5%<sub>(2)</sub> | 4%<sub>(6)</sub> | - |
| White/Caucasian | 2.5%<sub>(2)</sub> | 80%<sub>(121)</sub> | - |
| Asian/Asian American | 0 | 13%<sub>(19)</sub> | - |
| Hawaiian/Pacific Islander | 0 | <1%<sub>(1)</sub> | - |
| **APOL1 genotype (%)** |  |  |  |
| G0/G0 | 28%<sub>(20)</sub> | 100%<sub>(152)</sub> | - |
| G0/G1 | 26%<sub>(19)</sub> | 0 | - |
| G0/G2 | 15%<sub>(11)</sub> | 0 | - |
| G1/G1 | 10%<sub>(7)</sub> | 0 | - |
| G1/G2 | 15%<sub>(11)</sub> | 0 | - |
| G2/G2 | 6%<sub>(4)</sub> | 0 | - |

Lower eGFR in AA NEPTUNE participants could reflect social factors that negatively affect health [20]. If lower eGFR in AA primarily reflects social determinants, we reasoned it would be similar in AA participants with and without *APOL1* risk variants and lower than eGFR in noAA participants. Baseline eGFRs were stratified by ancestry (AA vs noAA) and eGFRs in AA participants further grouped by 0, 1, and 2 *APOL1* risk alleles (RA) (**Supplemental Table 1**). Median (IQR) eGFRs between groups were significantly different (p≤6×10^−6^) (**Figure 1**). Post-test, pairwise comparisons showed that the median eGFR ranks in noAA (n=140) and AA_0RA (n=20) participants were almost identical and significantly higher than mean eGFRs for both AA_2RA (n=22) and AA_1RA (n=30) participants. Together, these data suggest that in this cohort, the lower eGFR in AA is associated with *APOL1* risk alleles, but not with the social, physical, and institutional contexts shared by many AA.

**Figure 1:**
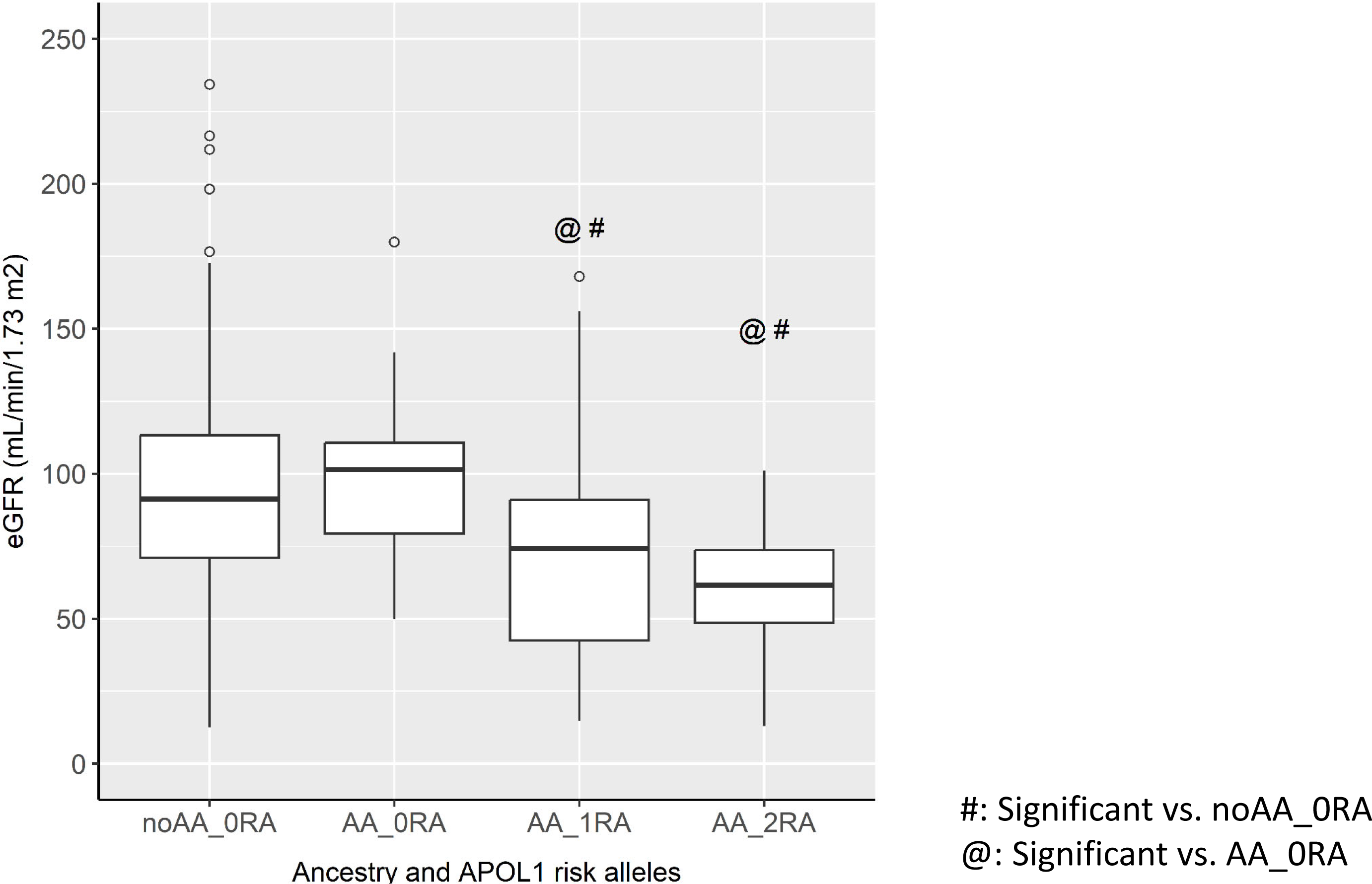
Boxplot of eGFR of the NEPTUNE study cohort participants. Individuals were stratified by inferred African ancestry (AA) or noAA (see Methods) and by number of *APOL1* risk alleles (RA) as 0RA, 1RA and 2RA. The Median (IQR) of eGFRs (ml/min/1.73 m^2^) were: noAA_0RA, 91 (71-113) [n=140]; AA_0RA, 101 (79-111) [n=20]; AA_1RA, 74 (43-91) [n=29] and AA_2RA, 62 (49-74) [n=21]. The median rank of these groups were significantly different (Kruskal Wallis, p≤2×10^−5^). Post-test comparisons were conducted using the Wilcoxon rank sum test. The median eGFR rank for noAA_0RA participants was significantly higher compared to the median eGFR ranks from AA_1RA (p≤0.01) and AA_2RA (p≤0.0002). The mean the eGFR rank from AA_0RA was significantly different to those from AA_1RA (p≤0.02) and AA_2RA (p≤0.002). There were no significant differences in the mean eGFR ranks between AA_0RA and noAA_0RA or between AA_1RA and AA_2RA. The p values were adjusted by the method of Benjamini & Hochberg. #, significantly different from noAA_0RA; @, significantly different from AA_0RA. Open circles represent outliers.

### Weighted gene co-expression analysis (WGCNA) and module-trait correlations

We used WGCNA to identify gene coexpression modules correlated with *APOL1* genotypes. Ancestry-specific patterns of genetic architecture and social factors can affect gene expression [21–24] and contribute to transcriptional heterogeneity [25]. To mitigate transcriptional heterogeneity reflecting ancestry and enrich for gene expression reflecting biologic pathways driving *APOL1* nephropathy, we created a WGCNA network using only individuals with AA. The resulting network comprises 42 gene coexpression modules clustered together into 12 higher order structures called metamodules (**Supplemental Figure 2**).

Next, we used the AA network to determine module-trait correlations in the whole NEPTUNE cohort (n=224) with the following traits: eGFR, log(1 + UPCR [urinary protein creatinine ratio]) (UPCR_lg), FSGS, AA, the number of *APOL1* risk alleles (*APOL1*_RA). A summary of the significant correlations is shown in **Figure 2**, while the Pearson correlation coefficients and nominal p-values are in **Supplemental Figure 3**. UPCR and eGFR are strong predictors of kidney disease progression [26]. In this analysis, eGFR and UPCR_lg concordantly correlated with multiple modules, indicating that the integrity of the glomerular filtration barrier is reflected in complex transcriptional regulation within the glomerulus. Interestingly, three modules, Darkturquoise, Salmon and Midnightblue, were significantly correlated with the number of *APOL1* risk alleles (p-Adj≤8×10^−4^, p-Adj≤1×10^−5^ and p-Adj≤2×10^−3^, respectively) as well as with FSGS. In addition, the Salmon module also correlated with AA (p-Adj≤0.042), which may suggest ancestry is a confounder of *APOL1* risk allele variants. However, the significant association of the Salmon module with the number *APOL1* risk alleles persisted even when module-trait correlations were conducted solely in AA participants (p-Adj≤0.029), indicating that the latter is not the case.

**Figure 2:**
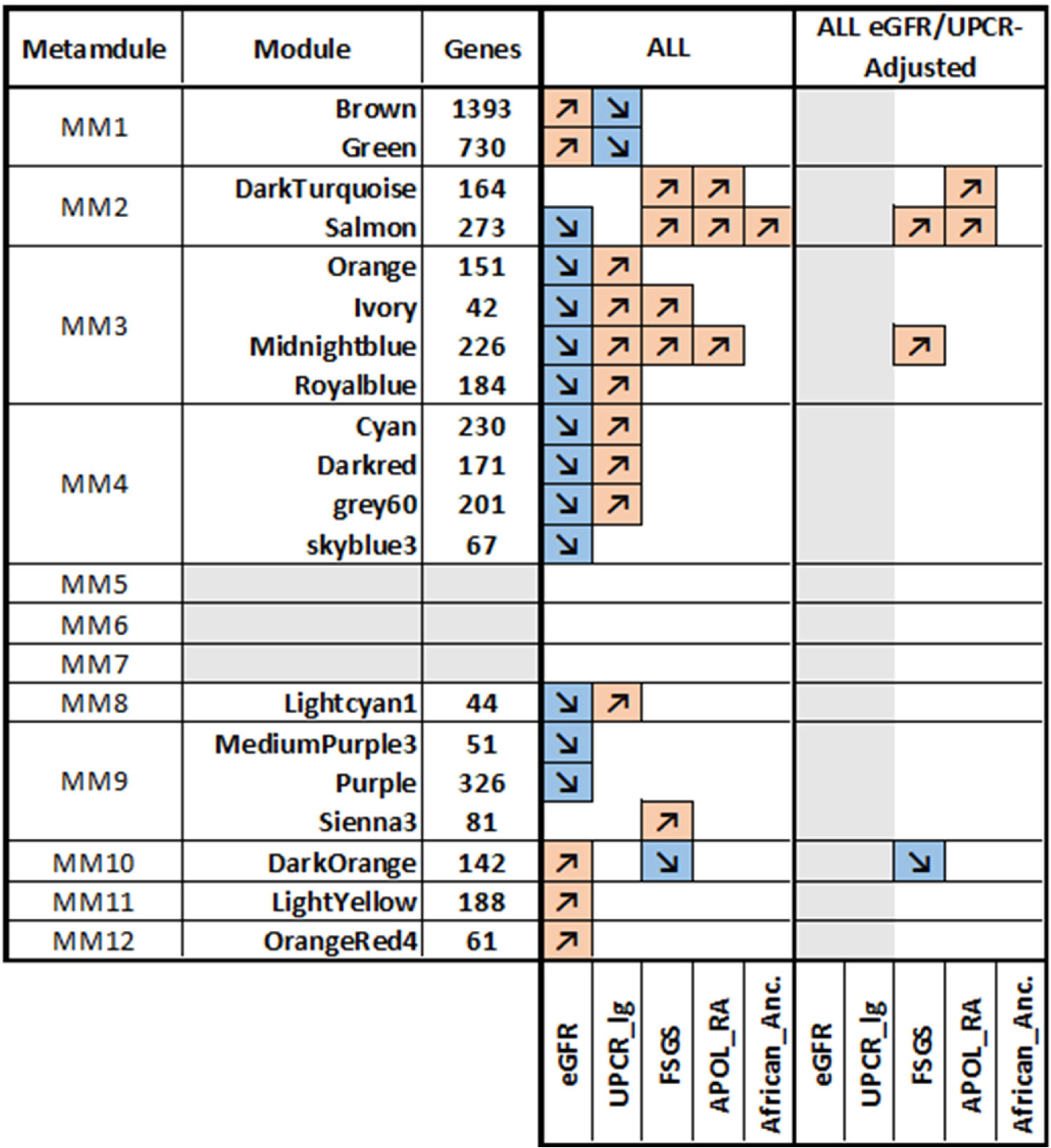
Module-Trait Correlations (MTC) analyses using the African ancestry network. Independent MTC were conducted in all NEPTUNE participants (ALL), and ALL after adjustment for eGFR and UPCR (ALL eGFR/UPCR-Adjusted). The following traits were analyzed: eGFR, log(1 + UPCR) (UPCR_lg), FSGS, *APOL1* risk alleles number (*APOL1*_RA), and inferred African ancestry (African_Anc). Multiple metamodules significantly associated with eGFR and UPCR_lg. In addition, 6 modules associated with FSGS, 3 of which, Darkturquoise, Salmon and Midnightblue, also significantly associated with the number of *APOL1* risk alleles (See **Supplemental Table 2** for the genes contained in these modules). Orange/Up-arrows represent positive correlations, while Blue/Down-arrows represent negative correlations. Only significant correlations are shown. For each trait, we adjusted the p-values using the Bonferroni correction to account for the multiple tests conducted (1 test for each module). The complete statistics and nominal p values are presented in **Supplemental Figure 3**. Grey areas represent null data *i.e.* the correlations with eGFR or UPCR_lg in the adjusted data; and lack of any significant correlation in modules within MM5, MM6 or MM7.

In addition to the number of *APOL1* risk alleles, the Darkturquoise, Salmon, and Midnightblue modules showed significant correlations with eGFR and UPCR_lg. In the NEPTUNE cohort, participants carrying 1 or 2 *APOL1* risk alleles had significantly lower eGFR compared to those homozygous for the G0 allele (**Figure 1**), while AA had marginally lower UPCR at the time of biopsy (**Table 1**). To account for this, we repeated the module-trait correlation analysis using eGFR- and UPCR-adjusted transcriptomes. Only the Darkturquoise and Salmon modules remained associated with the number of *APOL1* risk alleles, both with p-Adj≤0.008 (**Figure 2**). Darkturquoise and Salmon are the only modules clustered together in MM2 (**Supplemental Figure 2**); therefore, we focused on analyzing MM2 as a whole in subsequent analyses.

### Clinical outcomes associate with MM2 gene activation scores

To assess the clinical significance of the MM2 gene set (n=437 genes), we generated a mean z-score from the transcriptional data for each participant, who were then binned by tertiles of mean gene activation. Clinical outcomes were available on 191 NEPTUNE participants (60 AA; 131 noAA). An unadjusted Kaplan-Meier analysis using a log rank test was used to determine significant differences in outcomes in AA and noAA, respectively. AA with the highest MM2 tertile of gene activation scores had longer times to remission (p = 0.014), and trended to poorer outcomes, as captured by of the composite endpoint of kidney failure or loss of 40% of eGFR. This association and trend were not present in the noAA cohort (**Figure 3**). The most highly significant Molecular Signatures Database (MSigDB) Hallmark gene sets enrichments in MM2 were Epithelial Mesenchymal Transition (EMT), Estrogen Response Early and Apical Junction.

**Figure 3:**
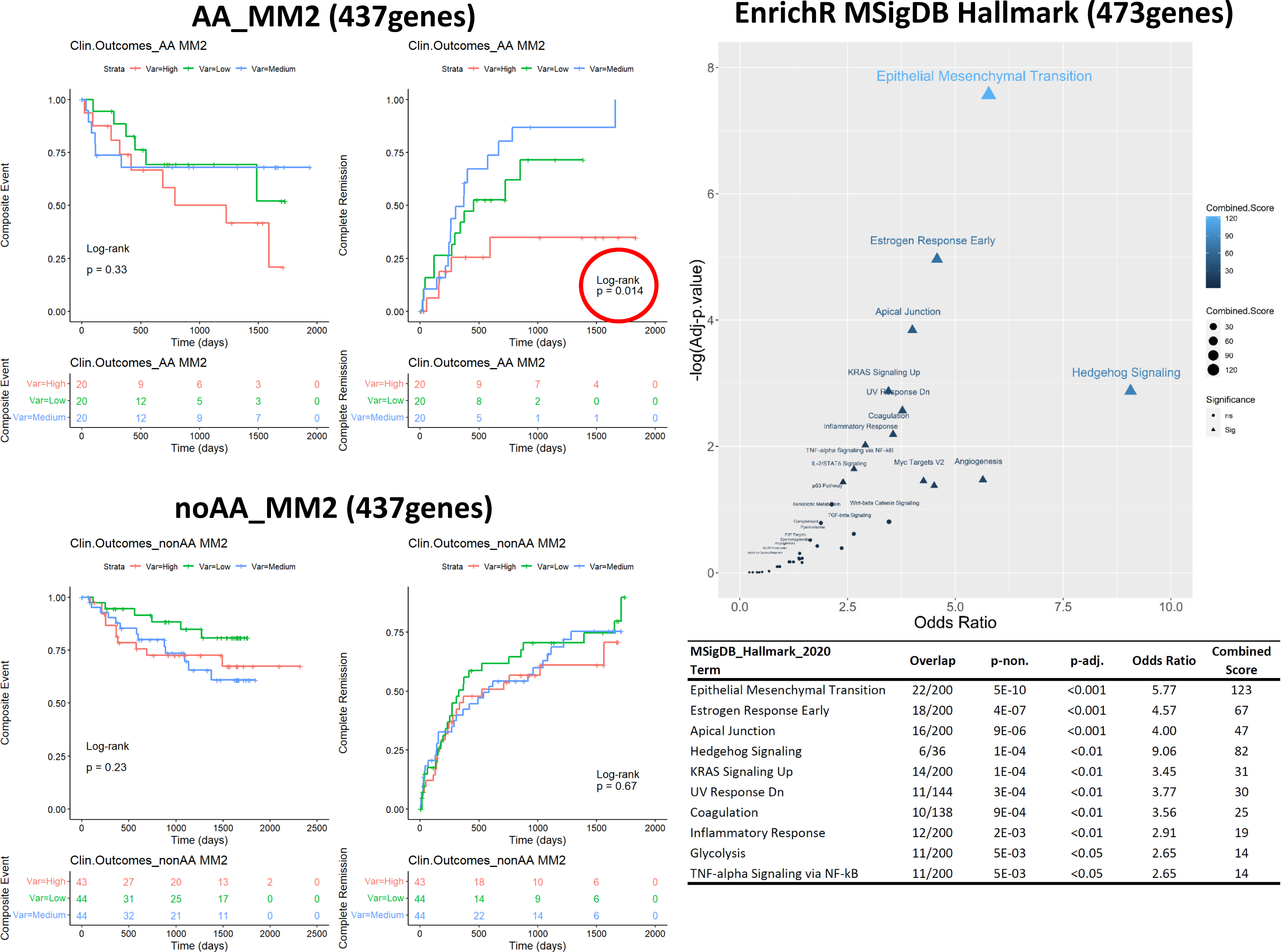
Kaplan-Meier curves stratified by tertiles of metamodule 2 (MM2) gene activation scores in NEPTUNE AA and noAA participants. Outcomes were time since kidney biopsy to a “Composite Event” of kidney failure or loss of 40% of eGFR, or to “Complete Remission” of proteinuria. Enrichr was used to obtain gene set enrichment analysis using the MSigDB Hallmark 2020 gene set (right panel and table).

In contrast to MM2, the Midnightblue module (n=226 genes) remained correlated with FSGS but was no longer associated with *APOL1* risk allele number after adjustment for UPCR_lg and eGFR in WGCNA analysis (**Figure 2**). Both AA and noAA with the highest tertile of Midnightblue gene activation-scores had significantly longer times to remission (AA, p≤0.008; noAA, p≤0.036), while only in AA the highest tertile presented a marginally significant faster decline in kidney function (p≤0.059) (**Supplemental Figure 4**). These data indicate that the Midnightblue gene module regulates the mechanisms resulting in FSGS histopathology, although the pathways may be modified by *APOL1*. The most highly enriched pathways in the Midnightblue module were TNF-alpha Signaling via NF-kB, EMT and Inflammatory Response (**Supplemental Figure 4**).

### Characteristic Direction (ChDir) identifies an *APOL1*-associated gene expression signature

To identify differentially expressed genes associated with *APOL1* risk alleles, we used a linear classifier called ChDir, which ranks genes by their contribution to the overall differences in expression between two classes [27]. To mitigate transcriptional heterogeneity reflecting ancestry, we generated a genome-wide ChDir signature by contrasting AA_2RA to AA_0RA participants. The top 1481 genes in the resulting ranked list (∼10% of the gene space) accounted for 75% of the gene expression differences (**Supplemental Table 3**). Next, the significance of the expression differences of the 1481 ChDir signature genes across different *APOL1* genotypes, we conducted permutational multivariate analysis of variance (PERMANOVA). We showed that AA_2RA and AA_0RA patients could be significantly separated (p≤0.001) using the Euclidean distance matrix of the 1481 ChDir gene space; and that in this context, the presence of *APOL1* risk alleles accounts for 6.4% of the total variability in gene expression between groups (**Figure 4-A**). Next, we repeated the analysis including all NEPTUNE participants stratified by inferred ancestry and number of *APOL1* risk alleles *i.e.* AA_2RA, AA_1RA, AA_0RA and noAA_0RA. Again, PERMANOVA shown significant separation of these groups (p≤0.001), with the presence of *APOL1* risk alleles accounting for 3.0% of the total variability (**Figure 4-B**). Post-test pairwise comparisons showed that the centroid of noAA was significantly separated from the centroids of AA_1RA (p≤0.006) and AA_2RA (p≤0.006). Similarly, the centroid from AA_0RA was significantly separated from those of AA_1RA (p≤0.024) and AA_2RA (p≤0.006). Importantly, the centroids of AA_0RA and noAA_0RA did not segregated (p≤0.25), nor did those from AA_1RA and AA_2RA (p≤0.46). As a negative control, we repeated the analysis using the lower 13151 ChDir signature genes. Within this gene space, PERMANOVA failed to significantly separate groups (**Supplemental Figure 5**). Finally, we determined if the *APOL1* ChDir gene signature could cluster an orthogonal dataset, glomerular transcriptomes from dual transgenic mice with the Tg26 transgene to model of HIV-associated nephropathy and with either *APOL1*-G0 (n=8) or *APOL1*-G2 (n=12) transgenes in podocytes [28, 29]. *APOL1* variants are strongly associated with HIV-associated nephropathy and FSGS. We identified 1237 mouse orthologues of the human 1481 *APOL1* ChDir signature genes, which significantly separated G2 from G0 mice (p≤0.032) accounting for 8.8% of the intergroup variance (**Figure 4-C**).

**Figure 4:**
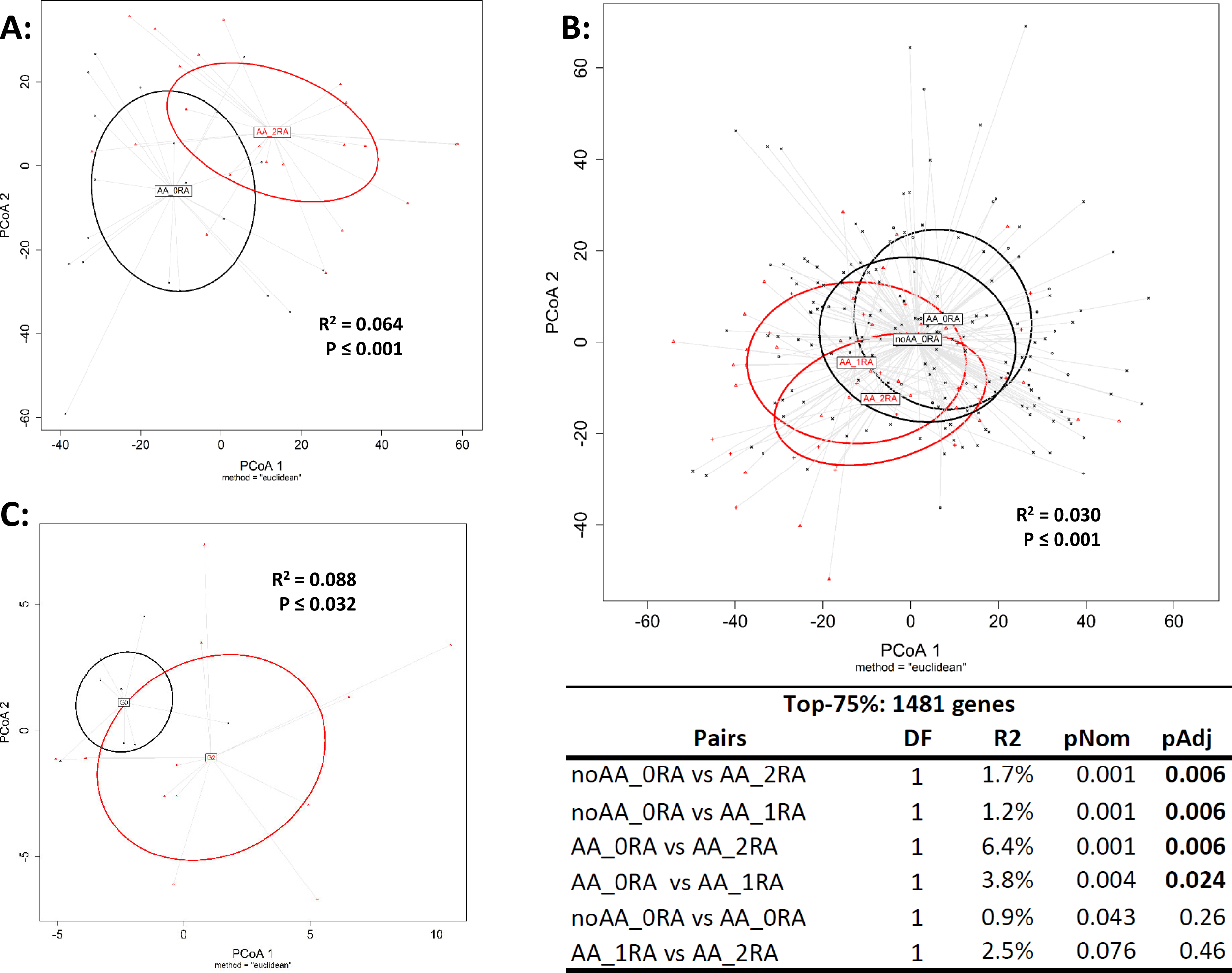
Principal Coordinates Analysis (PCoA) of the Euclidean distance matrix of the *APOL1* ChDir gene expression signature. Statistical analysis was performed using PERMANOVA. **Panel A** displays the first two PCoA coordinates, showing the separation between the centroids of the AA_0RA (black) and AA_2RA (red) groups, with the ellipses representing the 95% confidence intervals. PERMANOVA indicates a statistically significant separation between these two groups, as evidenced by R² = 0.064 and p ≤ 0.001. **Panel B** expands the analysis to four groups of NEPTUNE participants: noAA_0RA (black), AA_0RA (black), AA_1RA (red), and AA_2RA (red). The ellipses indicate the 95% confidence intervals for each group, and PERMANOVA confirms significant differences between the group centroids (R² = 0.030, p ≤ 0.001). Post-hoc pairwise comparisons (shown in the table) reveal significant separations between noAA_0RA and AA_1RA (p ≤ 0.006), as well as AA_0RA and AA_2RA (p ≤ 0.006), but no significant differences between noAA_0RA and AA_0RA (p = 0.26) or AA_1RA and AA_2RA (p = 0.46). **Panel C** illustrates the separation between transgenic mice expressing the APOL1-G0 (black) and APOL1-G2 (red) variants, using 1237 orthologues of the APOL1 ChDir gene signature. The analysis shows a significant separation between the two mice strains (R² = 0.088, p ≤ 0.032).

### Clinical outcomes associate with the ChDir *APOL1* gene signature activation scores

We generated z-scores for the 1481 genes in the *APOL1* gene signature for both AA and noAA NEPTUNE participants and stratified them into gene activation tertiles. Individuals with the highest gene activation scores had significantly faster times to the composite event in AA (p≤0.013) but not in noAA (p≤0.89). No significant differences were observed in either group for achieving complete remission (**Figure 5**). The most highly enriched pathways included Allograft Rejection, EMT, Inflammatory Response, KRAS Signaling Up and IL-6/JAK/STAT3 Signaling.

**Figure 5:**
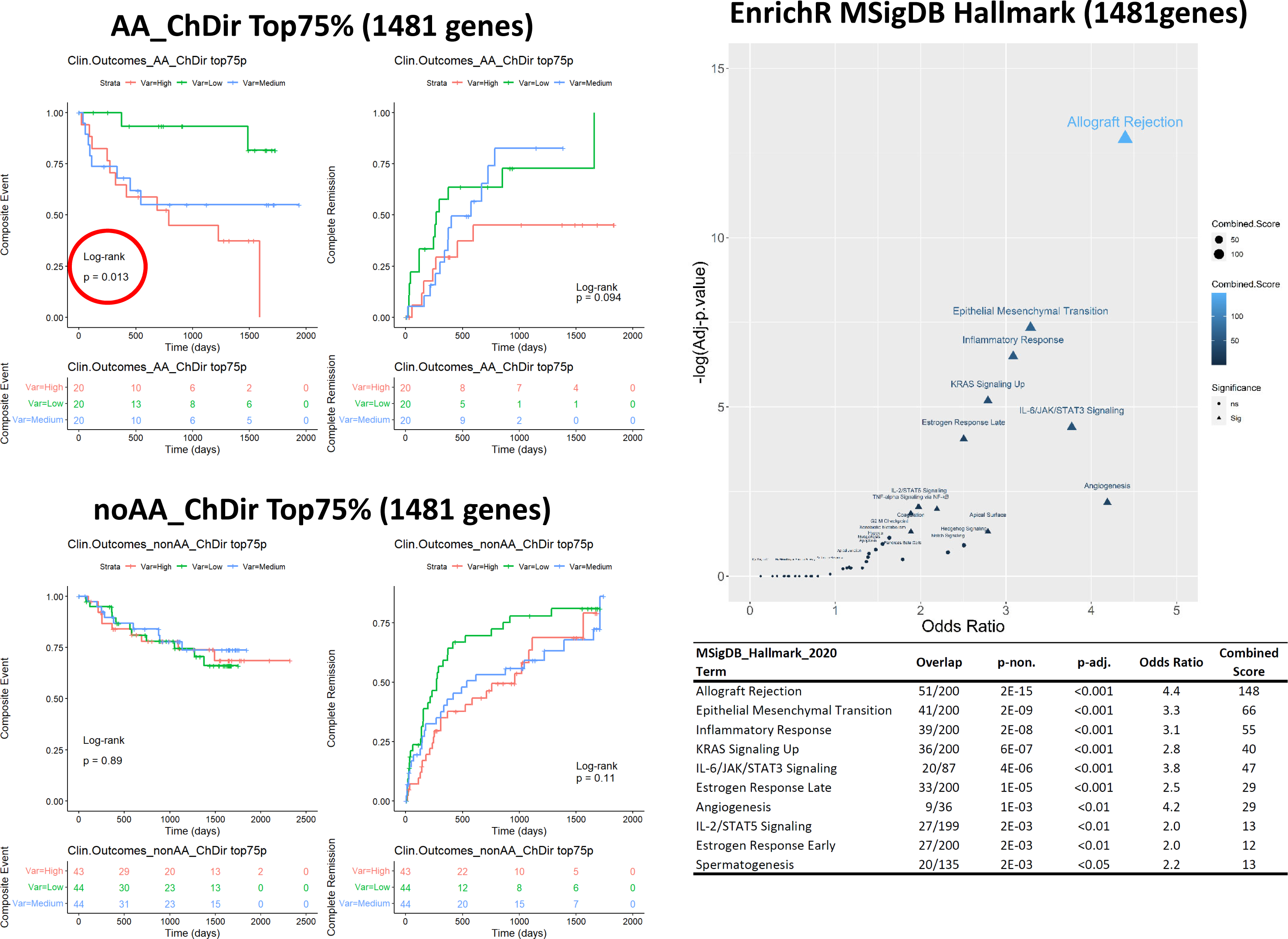
Kaplan-Meier curves, stratified by tertiles of the *APOL1* ChDir gene signature activation (n=1481 genes) in NEPTUNE AA and noAA participants. Outcomes were time since kidney biopsy to a “Composite Event” of kidney failure or loss of 40% of eGFR, or to “Complete Remission”. Enrichr was used to obtain gene set enrichment analysis using the MSigDB Hallmark 2020 gene set (right panel and table).

The enrichment terms from the ChDir *APOL1* gene signature and MM2, which correlated with *APOL1* risk allele number, were similar. Thus, we reasoned that the ChDir signature and MM2 gene set would overlap and be enriched with genes associated with *APOL1* pathogenesis. In the MM2_Salmon module 129 genes, from a total of 273 significantly overlapped with the 1481 genes of the *APOL1* signature (Adjusted Fisher exact test p≤4×10^−49^), while in the MM2_Darkturquoise module 62 genes, from a total of 164 significantly overlapped with the *APOL1* signature gene space (adjusted Fisher exact test p≤1×10^−19^). (**Supplementary Table 3**).

### Glomerular cell-identity gene signatures cluster *APOL1* genotypes

We next leveraged gene expression data from the Kidney Precision Medicine Project (KPMP) Kidney Tissue Atlas to generate cell-identity gene signatures containing genes enriched in, but not unique to, specific glomerular cell types (**Supplemental Table 4**). The glomerular cell identity signatures contained the following numbers of genes for each cell type: 1) glomerular visceral epithelium (POD, podocytes) 1814 genes, 2) glomerular parietal epithelium (PEC) 1220 genes, 3) glomerular capillary endothelium (EC-GC) 1298 genes, and 4) glomerular mesangium (MC) 1131 genes. In addition, we obtained cell-exclusive gene signatures containing genes with a positive fold change in only one kidney cell type (**Supplemental Table 5**).

As podocyte dysfunction characterizes primary nephrotic syndrome and podocytes express *APOL1* [30], we hypothesized that the transcriptional impact of *APOL1* variants would be primarily reflected in podocytes, in particular the podocyte identity gene signature. Using PERMANOVA, we next tested if the Euclidean distance matrix calculated from different glomerular cell-identity gene signatures would separate the NEPTUNE cohort by *APOL1* risk allele number. The distance matrix from the 1814 POD gene identity signature genes separated *APOL1* RA carriers from individuals with no *APOL1* RA, explaining 2% of the overall variability (p≤0.024, **Figure 6-A**). However, post-test comparisons showed that only transcriptomes from AA_2RA and noAA_0RA were significantly different (p-Adj≤0.048) using the POD-identity gene space. In addition to podocytes, the PEC identity signature was also able to significantly cluster patients (p≤0.001) by ancestry and presence of *APOL1* RA, explaining 3% of the overall variability (**Figure 6-B**). Post-test pairwise comparisons showed that the centroids from AA and noAA without *APOL1* risk alleles were significantly different from those from AA, who carried one or two *APOL1* risk alleles. In addition, the centroids from AA_0RA and noAA_0RA or AA_1RA and AA_2RA were not significantly different. Finally, the MC and ECGC identity gene signatures failed to cluster NEPTUNE participants by *APOL1* risk allele status (**Supplemental Figure 6**). Together, these results indicate that the impact of *APOL1* variants on glomerular cells transcriptomes was not limited to podocytes as we originally thought, but more markedly present in PECs.

**Figure 6.**
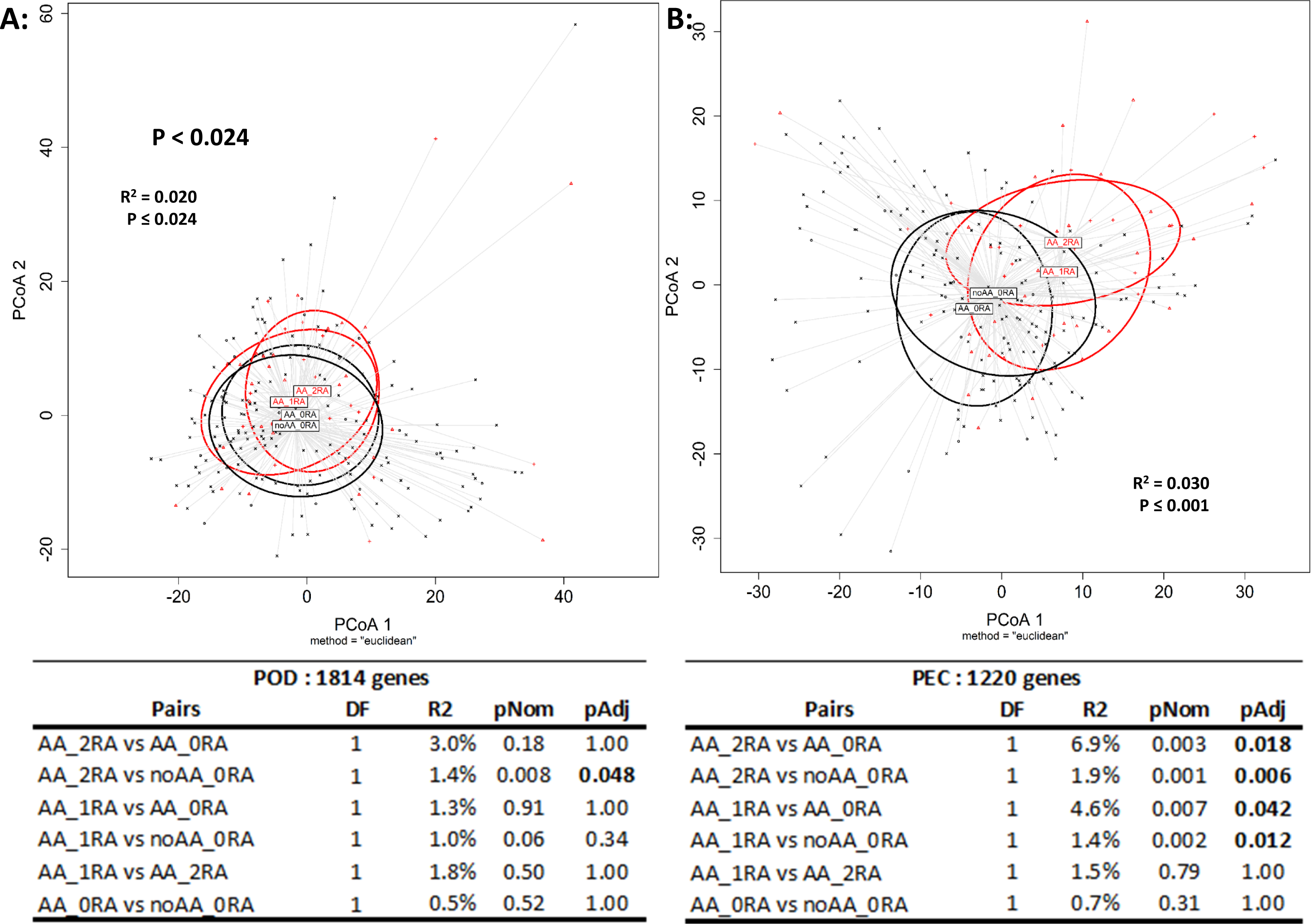
Principal Coordinate Analysis (PCoA) using the Euclidean distance matrices of podocyte (POD, 1814 genes) and parietal epithelial cell (PEC, 1220 genes) identity gene signatures. **Panel A** shows the two first coordinates and the separation of the centroids from noAA_0RA, AA_0RA, AA_1RA and AA_2RA NEPTUNE participants using the POD identity gene signature distance matrix. PERMANOVA analysis indicates that the centroids of these four groups are significantly separated (p≤0.024). Post-test comparisons show that only the noAA_0RA and AA_2RA centroids are statistically different in this gene space (p≤0.048). **Panel B** shows the first two coordinates of the same groups in the PEC identity signature gene distance matrix. PERMANOVA showed the group centroids were significantly separated (p≤0.001). Post-test pairwise comparisons showed that the centroid from noAA_0RA was significantly different from those from AA_1RA (p≤0.012) and AA_2RA (p≤0.006). Similarly, the centroid from AA_0RA was different from those from AA_1RA (p≤0.042) and AA_2RA (p≤0.018). Most importantly, the centroids from AA_0RA and noAA_0RA or AA_1RA and AA_2RA were not significantly separated. Red ellipses indicate patients with APOL1 risk alleles *i.e.* AA_2RA and AA_1RA, while black indicates AA_0RA and noAA_0RA.

Since the POD- and PEC-identity genes separated individuals by *APOL1* RA status, we hypothesized that these gene sets would by enriched in MM2. Thus, we used the Fisher exact test to evaluate the overlap between metamodules and cell-exclusive gene signatures (**Figure 7-A**). Our results indicate that MM2 (Salmon and Darkturquoise), which is associated with the number of *APOL1* risk alleles, reflects transcriptional programs active in PECs. On the other hand, MM4 (Cyan, Darkred, Grey60 and Skyblue3), and MM10 (DarkOrange), which associated with GFR and proteinuria (**Figure 2**), overlap with transcriptional programs in podocytes. The complete mapping of sub-region exclusive genes to metamodules can be found in **Supplemental Table 4**. Finally, we calculated the scores of MM2, MM4 and MM10 using a reference adult human kidney (GSE169285) dataset containing integrated single-cell and – nuclei transcriptomes. In this analysis, we provided the gene spaces of each of the aforementioned metamodules, to calculate an aggregate expression score across glomerular cells confirming that MM2 genes are most highly express in PECs, while MM4 and MM10 genes are most highly expressed in POD (**Figure 7-B**), confirming previous results.

**Figure 7-A:**
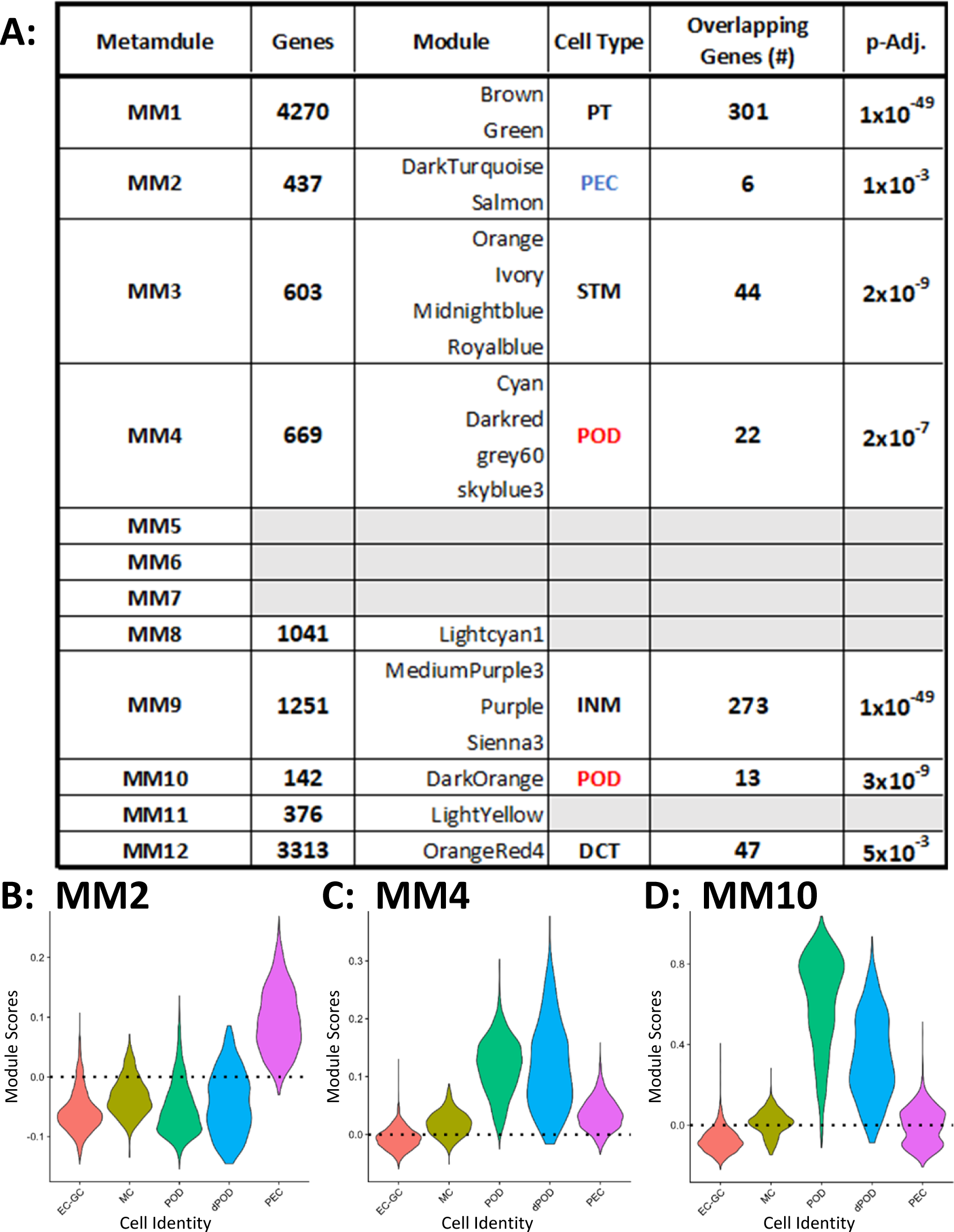
Overlap analysis between genes in metamodules and cell exclusive gene signatures obtained from the Kidney Precision Medicine Project. The total number of genes in each metamodule are presented in the “Genes” column. The number of cell-exclusive genes were as follows: POD, visceral epithelial cells (125 exclusive genes); PEC, parietal epithelial cells (26 exclusive genes); PT, proximal tubule cells (425 exclusive genes); DCT, distal convolute tubule cells (131 exclusive genes); SC, stromal cells (365 exclusive genes); and IMN, immune cells (950 exclusive genes). Adjusted-Fisher exact test p-values are presented in the “p-Adj” column. Only, significant enrichments are presented in the table. Expression scores of MM2 **(Panel B)**, MM4 **(Panel C)**, and MM10 **(Panel D)** in glomerular cells of the adult human kidney. The gene spaces for each metamodule (437, 669 and 142 genes, respectively) were provided to compute aggregate expression scores across glomerular cell populations. Glomerular capillary endothelium (EC-GC), glomerular mesangium (MC).

## DISCUSSION

Previous studies in the NEPTUNE cohort assessing the impact of *APOL1* kidney risk variants on glomerular transcriptomes, only included self-identified ‘black/African American’ participants diagnosed with FSGS [14, 31]. In this study, we analyzed the entire NEPTUNE cohort with glomerular RNAseq and *APOL1* genotype data, since *APOL1* kidney risk variants associate with steroid resistant nephrotic syndrome and worse kidney outcomes in people with membranous nephropathy [16–19]. Using two distinct analytic methods, WGCNA and ChDir, we identified glomerular gene signatures that significantly associated with *APOL1* risk allele number. These signatures shared 191 genes, and were associated with kidney outcomes in Black NEPTUNE participants. Both signatures were enriched in MSigDB Hallmark gene sets that represent EMT and inflammation pathways, but not cell death. The ChDir gene signature was validated with an orthogonal glomerular transcriptome dataset from mice with HIV-associated nephropathy that carry *APOL1* G0 or G2 transgenes [28, 29]. Using the KPMP expression data to generate kidney cell-identity gene signatures and gene module-scores in glomerular cells, we discovered that *APOL1* kidney risk variants not only modify podocyte transcriptomes but also impact the gene expression landscape of PECs. Collectively, these data suggest that *APOL1* kidney disease risk variants alter the podocyte cell state, leading to the activation of PECs, a cell strongly implicated in glomerular injury, through a yet to be defined paracrine pathway. Podocytes are specialized cells with epithelial and mesenchymal characteristics and constitutively synthesize *APOL1* [30, 32, 33]. The enrichment of genes from a PEC identity signature in the WGCNA metamodule that correlated with *APOL1* variant number was unexpected. However, podocytes and PECs share a common epithelial lineage during development until they diverge late in glomerulogenesis [34]. A growing body of evidence demonstrates that communication between podocytes and PECs mediates the onset and progression of podocytopathies [35, 36]. Genetic cell lineage tracing in transgenic mice has definitively shown that PEC activation after podocyte injury is a key driver of glomerular scarring and collapse [37]. Histologic analyses of human FSGS using cell marker studies demonstrate findings consistent with these mouse data [38, 39]. Multiple studies demonstrate a stereotypical response PECs to some types of podocyte injury, which is initiated by PEC proliferation, followed by PEC migration onto the glomerular tuft and culminating in deposition of extracellular matrix [34, 35, 40]. Activated PECs are identified specifically in regions of podocyte loss with fibrous synechiae between the capillary tufts and Bowman’s capsule [41].

The mechanisms of PEC activation by podocyte injury are just being defined and focus primarily on podocyte loss, although paracrine signaling between PECs and podocytes has been demonstrated [42–44]. Variant APOL1s associate with a spectrum of kidney disease phenotypes [4, 45], which include podocyte injury that could result in PEC differentiation. Multiple mechanisms have been proposed for *APOL1* podocytopathy, mostly focused on cytotoxicity especially in high interferon inflammatory states. However, IFNγ treatment drives organoid podocytes to a more immature phenotype with maintenance of junctional complexes with the slit diaphragm protein ZO1 and without podocyte loss [46]. Trajectory inference of G1 organoid podocytes showed a shift to more immature phenotypes [13], and data from other cell models suggest variant *APOL1* generate podocyte dedifferentiation phenotypes characteristic of podocytopathy [6, 47, 48]. Interestingly, two reports demonstrate *APOL1* expression in PECs in HIV- and COVID associated nephropathy [49, 50], raising the possibility that *APOL1* could directly regulate PEC activation in disease.

Both our WGCNA and ChDir analyses suggests that a single *APOL1* risk variant impacts the glomerular transcriptional landscape. Our data is consistent with a model whereby a single *APOL1* risk variant generates a transcriptional prodromal state, which permits development of kidney disease in the presence of specific secondary stresses/conditions. While the presence of two *APOL1* risk alleles robustly associates with kidney disease phenotypes, several studies suggest that heterozygous carriers may also be susceptible to kidney diseases [51–54]. A recent study involving a large West African cohort of Stage 2 through 5 CKD found that while biallelic *APOL1* risk variant carriers have 25% higher odds of developing CKD, participants carrying 1 risk allele present 18% higher CKD odds than homozygous G0 [55]. In addition, genetic modifiers affect the pathogenicity of *APOL1* risk variants including the haplotype background of the G0 [2] and G2 [1], and stop-gain variant in *APOL3* (p.Q58*), which was associated with increased CKD risk in individuals with a single *APOL1* kidney risk allele [56]. Together these studies highlight the need to extend genotyping beyond G1 and G2 polymorphisms [57] to more precisely characterized kidney risk. This study has limitations. NEPTUNE is unique in its age range and diversity and a replication cohort is not available. Other cohorts of people who have kidney transcriptomes are not diverse or do not include patients with nephrotic syndrome. In addition, the sample size is limited to 72 NEPTUNE participants with AA. Multi-omic annotation of kidney tissue obtained from appropriately powered cohorts of people with and without *APOL1* kidney disease will certainly better define the driver cells and pathogenic mechanisms [58].

## METHODS

### NEPTUNE glomerular transcriptomes and clinical data

Glomerular transcriptomes (n=224) from microdissected glomeruli were obtained from the NEPTUNE consortium with their associated clinical data. A variable called “inferred African ancestry” (AA) was defined positive for those patients having an *APOL1* genotype different from G0/G0, and/or a self-reported race of “Black/African American” (n=72); while G0/G0 individuals, who did not self-identify as “Black/African American,” were defined as not having African ancestry (noAA, n=152). In some analyses, the gene count matrix was adjusted for eGFR and UPCR by fitting a linear model and extracting the residuals.

### *APOL1* transgenic mouse glomerular microarray data

We generated glomerular transcriptomes from 40-day old Tg26 transgenic mice with *APOL1*-G0 or *APOL1*-G2 transgenes controlled by the murine *Nphs1* promoter [28, 29] using the Mouse Gene 2.0 ST Array (Affymetrix).

### Weighted Gene Co-Expression Network Analysis (WGCNA) in individuals with AA

The 72 individuals with AA were selected to build a gene coexpression network with the R package WGCNA [59, 60]. The significance threshold in module-trait correlation analysis was adjusted by Bonferroni.

### Kaplan-Meier analysis with clinical outcomes

We conducted Kaplan-Meier analysis using gene expression z-scores tertiles as surrogates for the gene module activation as previously reported [61, 62]. The endpoint outcomes were “Complete Remission” or “Composite Event”. Differences between the tertiles [high, medium, low] curves were tested using the log-rank test. Independent analyses were conducted on AA and noAA. We analyzed the following gene spaces: 1) MM2 (437 genes), 2) Midnightblue module (226 genes) and 3) the *APOL1* ChDir signature (1481 genes).

### Functional enrichment analyses

We utilized EnrichR [63, 64] to conduct functional enrichment analysis using the Molecular Signatures Database (MSigDB) Hallmark gene sets [65, 66].

### Characteristic Direction (ChDir)

ChDir [27] is a geometric multivariate approach to differential gene expression and was previously used by us and other investigators to obtain transcriptional signatures from proximal tubules [67, 68], podocytes [69], and human kidney cancerous cells [70]. ChDir generates normalized gene vectors representing the fractional contribution of each gene to the overall transcriptional differences between classes, which allows the extraction of the top-scoring classifier genes accounting for 75% of discrimination between classes. In our analyses, these genes represent a signature that differentiates individual by number of *APOL1* kidney risk alleles.

### Single-cell (sc) and single-nucleus (sn) RNAseq matrices

We used sc- and snRNAseq transcriptomes from the KPMP to generate Cell-Identity and Cell-Exclusive gene signatures from 13 anatomical sub-regions of the kidney: 1) POD, 2) PEC, 3) MC, 4) EC-GC, 5) Proximal Tubule (PT), 6) Loop of Henle thin portion (Thin Limbs), 7) Loop of Henle thick portion (TAL), 8) Distal Convolution (DCT), 9) Connecting Tubule (CNT), 10) Collecting Duct (CD), 11) Endothelium Non-Glomerular (EC), 12) Stroma Non-Glomerular (SC), and 13) Immune (IMN). The total number of genes on each signature can be found in **Supplemental Table 5**.

### Permutational Multivariate Analysis of Variance (PERMANOVA) and Principal Coordinates Analysis (PCoA)

We used PERMANOVA [71, 72], as implemented in the R package ‘vegan’ [73], to determine the statistical significance of the ability of different gene sets to distinguish between two or more groups of patients or transgenic mice. We inputted the Euclidian distance matrix as it emphasizes the actual proximity of gene expression values [74] as opposed to WGCNA and ChDir analyses, which are leveraged towards covariance analysis. When more than two groups were compared, p values were corrected using the function *pairwise.adonis()* with default parameters [75]. PCoA was performed on the Euclidean distance matrix to visualize the separation between groups in a reduced dimensional space.

### Module Scores

Gene module scores for MM2, MM4, and MM10 were generated in glomerular cells using the dataset from the “Integrated Single-nucleus and Single-cell RNA-seq of the Adult Human Kidney” (GSE169285).

### Statistics

Unless otherwise noted, all data were analyzed with “R: A language and environment for statistical computing and graphics” (https://www.R-project.org/). Some data inspection, cleaning or formatting were conducted in Notepad++ (https://notepad-plus-plus.org/) or in Microsoft Excel, which was also used to prepare tables for publication.

### Study approval

The NEPTUNE protocol is approved by the University of Michigan IRBMED central IRB (HUM00158219), and the Cleveland Clinic IRB (15-182). The KPMP study protocol is approved using a central IRB at the Human Research Protection Office of Washington University in St. Louis (IRB no. 201902013). NEPTUNE and KPMP participants provided written informed consent prior to enrollment. All animal studies were conducted under oversight of the Case Western Reserve University’s Institutional Animal Care and Use Committee (Protocol 2012-0099).

## Data Availability

All data produced in the present study are available upon reasonable request to the authors

## AUTHOR CONTRIBUTIONS

A.G-V, J.F.O and J.R.S conceived and designed research;

Z.W, J.F.O, J.R.S and L.A.B. conducted experiments with the mice;

A.G-V and V.N curated the data;

A.G-V analyzed data;

A.G-V, J.F.O and J.R.S interpreted results;

A.G-V. and J.R.S wrote the manuscript and figures with input from all authors.

All authors provided critical feedback and helped shape the research, analysis and manuscript. All authors approved the final version of the manuscript.

## ACKNOWLEDGEMENTS

This work was supported by National Institute of Diabetes and Digestive and Kidney Diseases (NIDDK) grants K01DK128304, 5T32DK07470, R01DK097836 and R01DK108329. The NEPTUNE consortium, U54-DK083912, is a part of the NIH Rare Disease Clinical Research Network (RDCRN), supported through collaboration between the Office of Rare Diseases Research, National Center for Advancing Translational Sciences, and the NIDDK. See Supplemental Acknowledgments for NEPTUNE consortium details. KPMP is a multiyear project funded by the NIDDK with the purpose of understanding and finding new ways to treat chronic kidney disease and acute kidney injury. See Supplemental Acknowledgments for KPMP consortium details.

The Kidney Precision Medicine Project is funded by the following NIDDK grants: U01DK133081, U01DK133091, U01DK133092, U01DK133093, U01DK133095, U01DK133097, U01DK114866, U01DK114908, U01DK133090, U01DK133113, U01DK133766, U01DK133768, U01DK114907, U01DK114920, U01DK114923, U01DK114933, U24DK114886, UH3DK114926, UH3DK114861, UH3DK114915, and UH3DK114937.

We thank NEPTUNE and KPMP participants and their families, who contributed their time, biospecimens, clinical data and kidney tissue for research purposes and without whom this study would not be possible.

## Supplemental Materials

### SUPPLEMENTAL ACKNOWLEDGEMENTS

#### For NEPTUNE

The Nephrotic Syndrome Study Network (NEPTUNE) is part of the Rare Diseases Clinical Research Network (RDCRN), which is funded by the National Institutes of Health (NIH) and led by the National Center for Advancing Translational Sciences (NCATS) through its Division of Rare Diseases Research Innovation (DRDRI). NEPTUNE is funded under grant number U54DK083912 as a collaboration between NCATS and the National Institute of Diabetes and Digestive and Kidney Diseases (NIDDK). Additional funding and/or programmatic support is provided by the University of Michigan, NephCure Kidney International, Alport Syndrome Foundation, and the Halpin Foundation. RDCRN consortia are supported by the RDCRN Data Management and Coordinating Center (DMCC), funded by NCATS and the National Institute of Neurological Disorders and Stroke (NINDS) under U2CTR002818.

**Members of the Nephrotic Syndrome Study Network (NEPTUNE):**

**NEPTUNE Collaborating Sites**

*Atrium Health Levine Children’s Hospital, Charlotte, SC*: Susan Massengill*, Layla Lo^#^

*Cleveland Clinic, Cleveland, OH*: Katherine Dell*, John O’Toole*, John Sedor**, Victoria Grange^#^

*Children’s Hospital, Los Angeles, CA*: Ian Macumber*, Alyssa Parry^#^

*Children’s Mercy Hospital, Kansas City, MO*: Tarak Srivastava*, Kelsey Markus^#^

*Cohen Children’s Hospital, New Hyde Park, NY*: Christine Sethna*, Suzanne Vento^#^

*Columbia University, New York, NY:* Pietro Canetta*

*Duke University Medical Center, Durham, NC:* Opeyemi Olabisi*, Rasheed Gbadegesin**, Maurice Smith^#^

*Emory University, Atlanta, GA:* Laurence Greenbaum*, Chia-shi Wang*, Emily Yun^#^

*The Lundquist Institute, Torrance, CA:* Sharon Adler*, Janine LaPage^#^

*John H Stroger Cook County Hospital, Chicago, IL:* Amatur Amarah*

*Johns Hopkins Medicine, Baltimore, MD:* Meredith Atkinson*, Sara Boynton^#^

*Mayo Clinic, Rochester, MN:* John Lieske, Marie Hogan, Fernando Fervenza

*Medical University of South Carolina, Charleston, SC:* David Selewski*, Cheryl Alston^#^

*Montefiore Medical Center, Bronx, NY:* Kim Reidy*, Michael Ross*, Frederick Kaskel**, Patricia Flynn^#^

*New York University Medical Center, New York, NY:* Laura Malaga-Dieguez*, Olga Zhdanova**, Laura Jane Pehrson^#^, Melanie Miranda^#^

*The Ohio State University College of Medicine, Columbus, OH*: Salem Almaani*, Laci Roberts^#^

*Stanford University, Stanford, CA:* Richard Lafayette*, Shiktij Dave^#^

*Temple University, Philadelphia, PA:* Iris Lee**

*Texas Children’s Hospital at Baylor College of Medicine, Houston, TX*: Shweta Shah*, Sadaf Batla^#^ ^#^

*University Health Network Toronto:* Heather Reich*, Michelle Hladunewich**, Paul Ling^#^, Martin Romano^#^

*University of California at San Francisco, San Francisco, CA*: Paul Brakeman*, Daniel Schrader

*University of Colorado Anschutz Medical Campus, Aurora, CO*: James Dylewski* Nathan Rogers^#^

*University of Kansas Medical Center, Kansas City, KS*: Ellen McCarthy*, Catherine Creed^#^

*University of Miami, Miami, FL:* Alessia Fornoni*, Miguel Bandes^#^

*University of Michigan, Ann Arbor, MI:* Matthias Kretzler*, Laura Mariani*, Zubin Modi*, A Williams^#^, Roxy Ni^#^

*University of Minnesota, Minneapolis, MN:* Patrick Nachman*, Michelle Rheault*, Amy Hanson^#^, Nicolas Rauwolf^#^

*University of North Carolina, Chapel Hill, NC:* Vimal Derebail*, Keisha Gibson*, Anne Froment^#^, Mary Mac McGown Collie^#^

*University of Pennsylvania, Philadelphia, PA:* Lawrence Holzman*, Kevin Meyers**, Krishna Kallem^#^, Aliya Edwards^#^

*University of Texas San Antonio, San Antonio, TX*: Samin Sharma**

*University of Texas Southwestern, Dallas, TX:* Elizabeth Roehm*, Kamalanathan Sambandam**, Elizabeth Brown**, Jamie Hellewege

*University of Washington, Seattle, WA:* Ashley Jefferson*, Sangeeta Hingorani**, Katherine Tuttle^**§^, Linda Manahan ^#^, Emily Pao^#^, Kelli Kuykendall^§^

*Wake Forest University Baptist Health, Winston-Salem, NC:* Jen Jar Lin**

*Washington University in St. Louis, St. Louis, MO*: Vikas Dharnidharka*

**Data Analysis and Coordinating Center:** *University of Michigan:* Matthias Kretzler*, Brenda Gillespie**, Laura Mariani**, Zubin Modi**, Eloise Salmon**, Howard Trachtman**, Tina Mainieri, Gabrielle Alter, Michael Arbit, Hailey Desmond, Sean Eddy, Damian Fermin, Wenjun Ju, Maria Larkina, Chrysta Lienczewski, Rebecca Scherr, Jonathan Troost, Amanda Williams, Yan Zhai; *Arbor Collaborative for Health:* Colleen Kincaid, Shengqian Li, Shannon Li; *Cleveland Clinic:* Crystal Gadegbeku**, *Duke University:* Laura Barisoni**; John Sedor**, *Harvard University:* Matthew G Sampson**; *Northwestern University:* Abigail Smith**; *University of Pennsylvania:* Lawrence Holzman**, Jarcy Zee**

**Digital Pathology Committee:** Carmen Avila-Casado *(University Health Network)*, Serena Bagnasco *(Johns Hopkins University)*, Lihong Bu *(Mayo Clinic)*, Shelley Caltharp *(Emory University)*, Clarissa Cassol *(Arkana)*, Dawit Demeke *(University of Michigan)*, Brenda Gillespie *(University of Michigan)*, Jared Hassler *(Temple University)*, Leal Herlitz *(Cleveland Clinic)*, Stephen Hewitt *(National Cancer Institute)*, Jeff Hodgin *(University of Michigan)*, Danni Holanda *(Arkana)*, Neeraja Kambham *(Stanford University)*, Kevin Lemley, Laura Mariani *(University of Michigan)*, Nidia Messias *(Washington University)*, Alexei Mikhailov *(Wake Forest)*, Vanessa Moreno *(University of North Carolina)*, Behzad Najafian *(University of Washington)*, Matthew Palmer *(University of Pennsylvania)*, Avi Rosenberg *(Johns Hopkins University)*, Virginie Royal *(University of Montreal)*, Miroslav Sekulik *(Columbia University)*, Barry Stokes *(Columbia University)*, David Thomas *(Duke University)*, Ming Wu *(University of New York)*, Michifumi Yamashita *(Cedar Sinai)*, Hong Yin *(Emory University)*, Jarcy Zee *(University of Pennsylvania)*, Yiqin Zuo *(University of Miami)*. Co-Chairs: Laura Barisoni *(Duke University)*, Cynthia Nast *(Cedar Sinai)*.

#### For KPMP

*American Association of Kidney Patients, Tampa, FL*: Richard Knight

*Beth Israel Deaconess, Boston, MA*: Stewart Lecker, Isaac Stillman

*Brigham & Women’s Hospital, Boston, MA*: Gearoid McMahon, Sus Waikar, Astrid Weins

*Broad Institute, Cambridge, MA*: Nir Hacohen, Paul Hoover

*Case Western Reserve, Cleveland, OH*: Mark Aulisio, Will Bush, Dana Crawford

*Cleveland Clinic, Cleveland, OH*: Leslie Cooperman, Leal Herlitz, John O’Toole, Emilio Poggio, John Sedor

*Columbia University, New York, NY*: Paul Appelbaum, Olivia Balderes, Jonathan Barasch, Andrew Bomback, Vivette D’agati, Krzysztof Kiryluk, Karla Mehl, Ning (Sunny) Shang, Chenhua Weng

*Duke University, Durham, NC*: Laura Barisoni

*European Molecular Biology Laboratory, Heidelberg, Germany*: Theodore Alexandrov

*Indiana University, Indianapolis, IN*: Tarek Ashkar, Daria Barwinska, Pierre Dagher, Kenneth Dunn, Michael Eadon, Michael Ferkowicz, Katherine Kelly, Timothy Sutton, Seth Winfree

*John Hopkins University, Baltimore, MD*: Steven Menez, Chirag Parikh, Avi Rosenberg, Pam Villalobos

*Joslin Diabetes Center, Boston, MA*: Sylvia Rosas, Mark Williams

*Mount Sinai, New York, NY*: Evren Azeloglu, Cijang (John) He, Ravi Iyengar

*Ohio State University, Columbus, OH*: Samir Parikh

*Pacific Northwest National Laboratories, Richland, WA*: Chris Anderton, Ljiljana Pasa-Tolic, Dusan Velickovic

*Parkland Center for Clinical Innovation, Dallas, TX*: George (Holt) Oliver

*Patient Advocates*: Joseph Ardayfio, Jack Bebiak, Keith Brown, Taneisha Campbell, Catherine Campbell, Lynda Hayashi, Nichole Jefferson, Robert Koewler, Glenda Roberts, John Saul, Anna Shpigel, Edith Christine Stutzke, Lorenda Wright

*Princeton University, Princeton, NJ*: Rachel Sealfon, Olga Troyanskaya

*Providence Medical Research Center, Spokane, WA*: Katherine Tuttle *Stanford University, Palo Alto, CA*: Yury Goltsev

*University of California San Diego, La Jolla, CA*: Blue Lake, Kun Zhang

*University of California San Francisco, San Francisco, CA*: Dejan Dobi, Maria Joanes, Zoltan Laszik, Garry Nolan, Andrew Schroeder

*University of Michigan, Ann Arbor, MI*: Ulysses Balis, Oliver He, Jeffrey Hodgin, Matthias Kretzler, Laura Mariani, Rajasree Menon, Edgar Otto, Jennifer Schaub, Becky Steck

*University of Pittsburgh, Pittsburgh, PA*: Michele Elder, Daniel Hall, John Kellum, Mary Kruth, Raghav Murugan, Paul Palevsky, Parmjeet Randhawa, Matthew Rosengart, Sunny Sims-Lucas, Mary Stefanick, Stacy Stull, Mitchell Tublin

*University of Washington, Seattle, WA*: Charles Alpers, Ian De Boer, Malia Fullerton, Jonathan Himmelfarb, Robyn Mcclelland, Sean Mooney, Stuart Shankland, Kayleen Williams, Kristina Blank, Ashveena Dighe

*UT Health San Antonio, San Antonio, TX*: Kumar Sharma, Guanshi Zhang

*UT Southwestern Medical Center, Dallas, TX*: Susan Hedayati, Asra Kermani, Simon Lee, Christopher Lu, Tyler Miller, Orson Moe, Harold Park, Kamalanathan Sambandam, Francisco Sanchez, Jose Torrealba, Toto Robert, Miguel Vazquez, Nancy Wang

*Washington University in St. Louis, St. Louis, MO*: Joe Gaut, Sanjay Jain, Anitha Vijayan

*Yale University, New Haven, CT*: Tanima Arora, Randy Luciano, Dennis Moledina, Ugwuowo Ugochukwu, Francis Perry Wilson

## SUPPLEMENTAL FIGURES

**Supplemental Figure 1:**
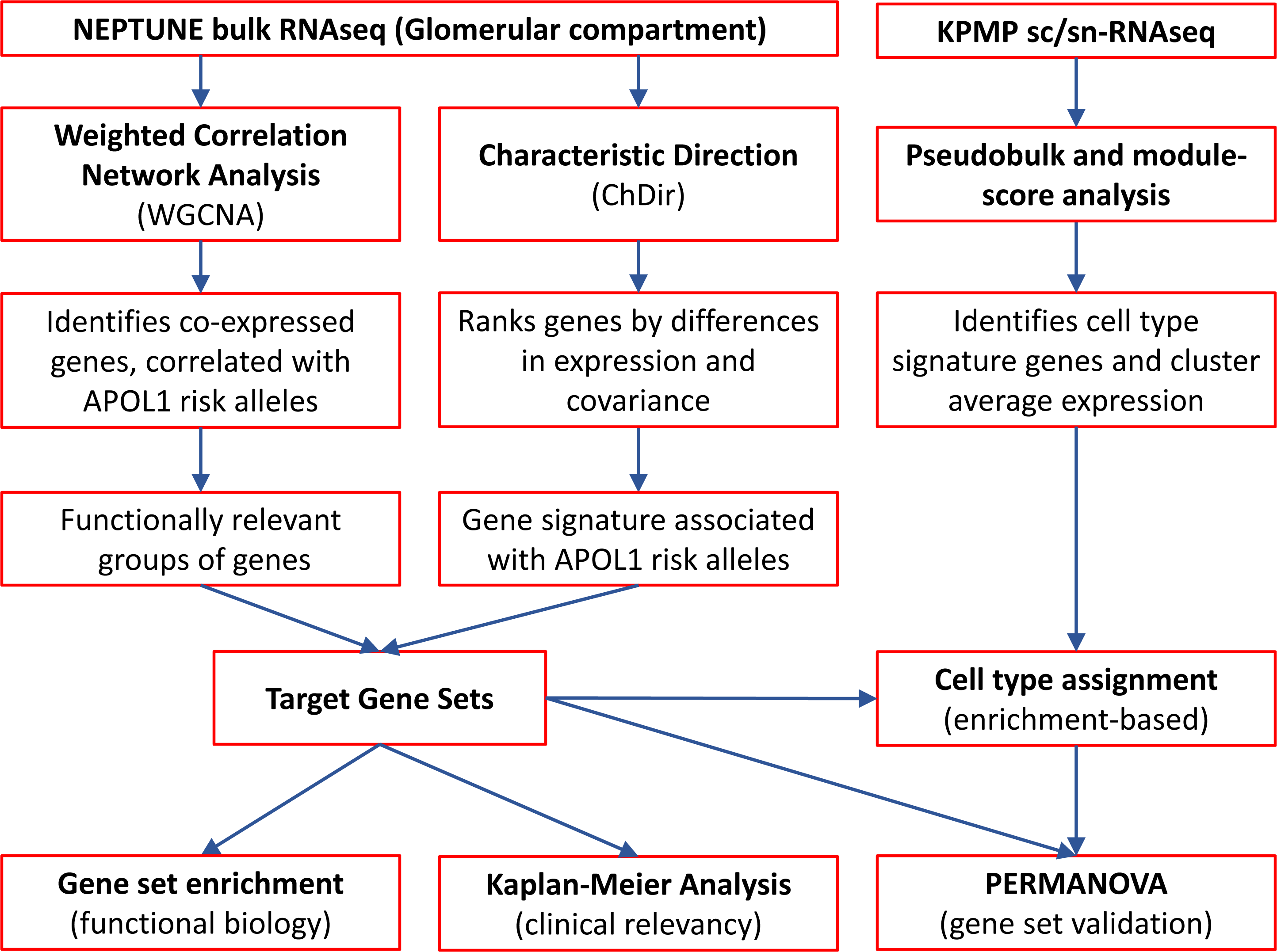
Overview of the methodology used in this manuscript.

**Supplemental Figure 2:**
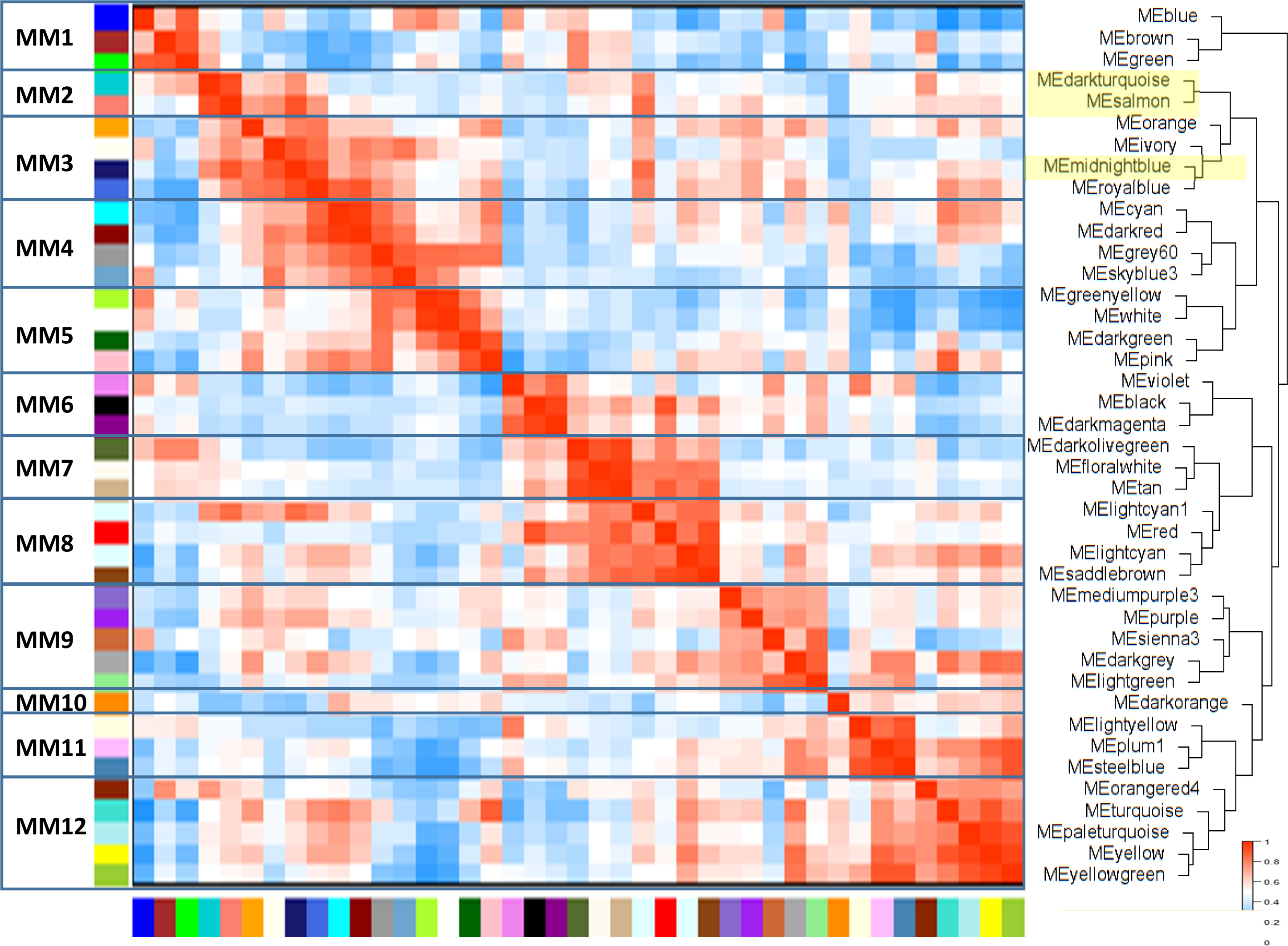
Module eigengene (ME) heatmap and dendrogram of the African ancestry (AA) network. Modules (shown on the right) of genes with correlated expression are grouped into metamodules (MM) shown on the left, which are derived from eigengene heatmap correlations and dendrogram structure. Some modules relevant in our downstream analyses are highlighted in yellow. The ME of the “Grey” module, containing one uncorrelated gene (*SRGAP3*) is not shown.

**Supplemental Figure 3:**
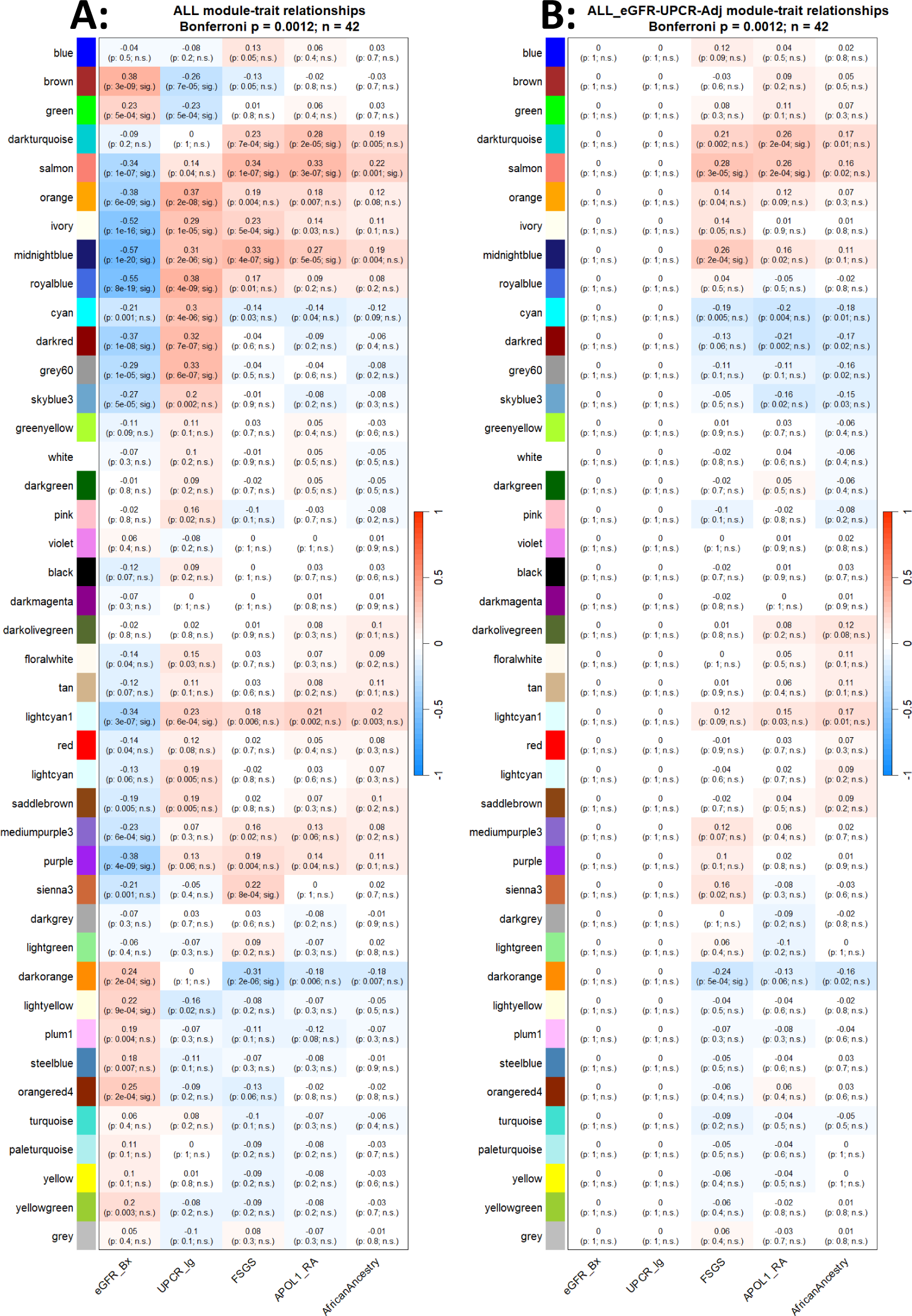
Module trait correlations for. Panel A) all participants (ALL), Panel B) ALL participants after adjusting by eGFR and UPCR (ALL_eGFR-UPCR-Adj). Each row corresponds to module eigengene and the rows are the traits. The values in the cells are Pearson correlation coefficient (r) and in parenthesis the nominal p-value, as well as the designation “sig.” if p≤0.012, the Bonferroni significant threshold or “n.s.” for p>0.012. The cells are color coded for the magnitude of r using the scale on the right (1, 0, −1; red, white, blue).

**Supplemental Figure 4:**
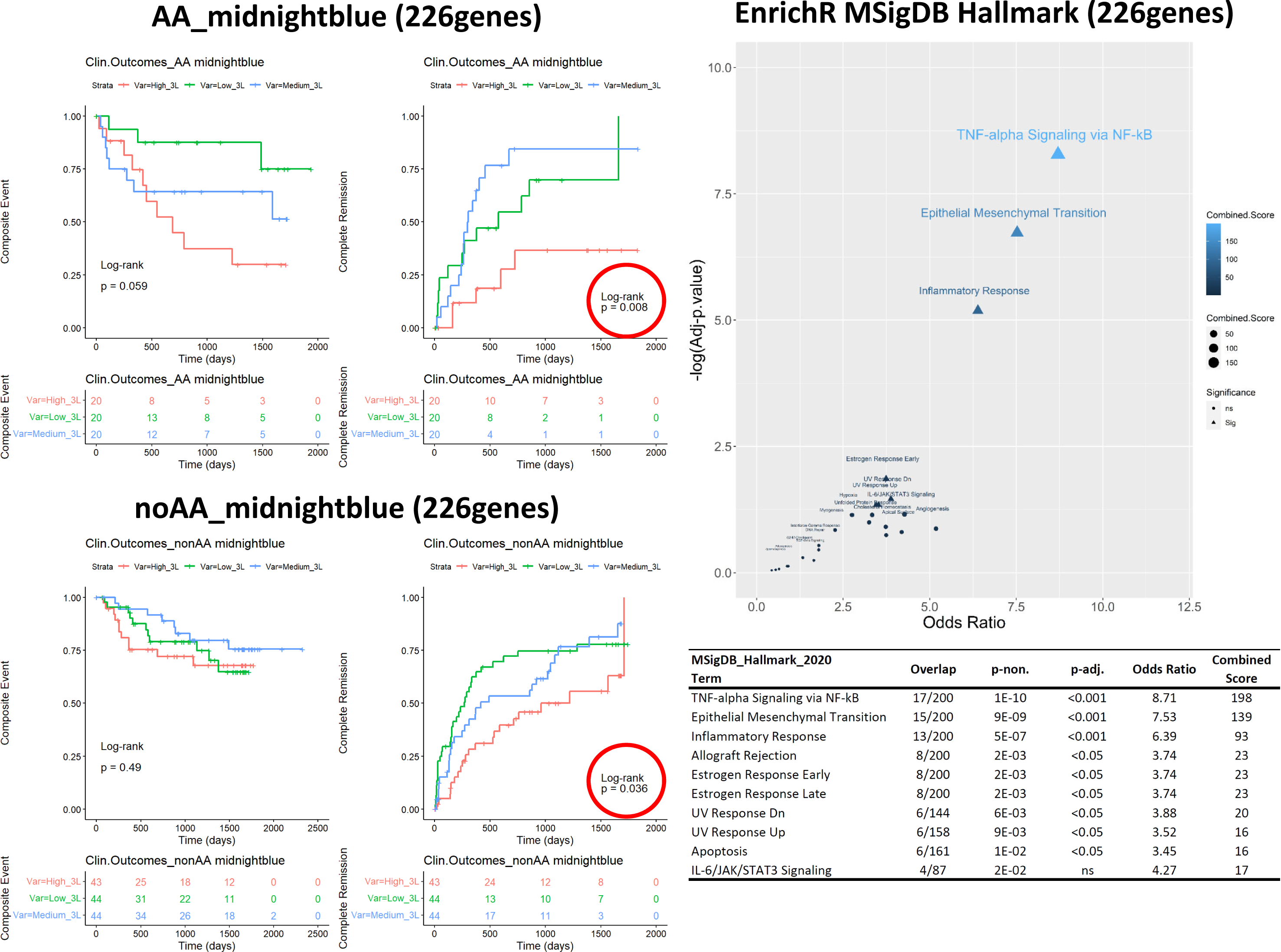
Unadjusted Kaplan-Meier curves stratified by tertiles of module Midnightblue (226 genes) gene activation scores in NEPTUNE AA and noAA participants. Outcomes were time since kidney biopsy to a “Composite Event” of kidney failure or loss of 40% of eGFR, or to “Complete Remission”. Enrichr was used to obtain gene set enrichment analysis using the MSigDB Hallmark 2020 gene set (right panel and table).

**Supplemental Figure 5:**
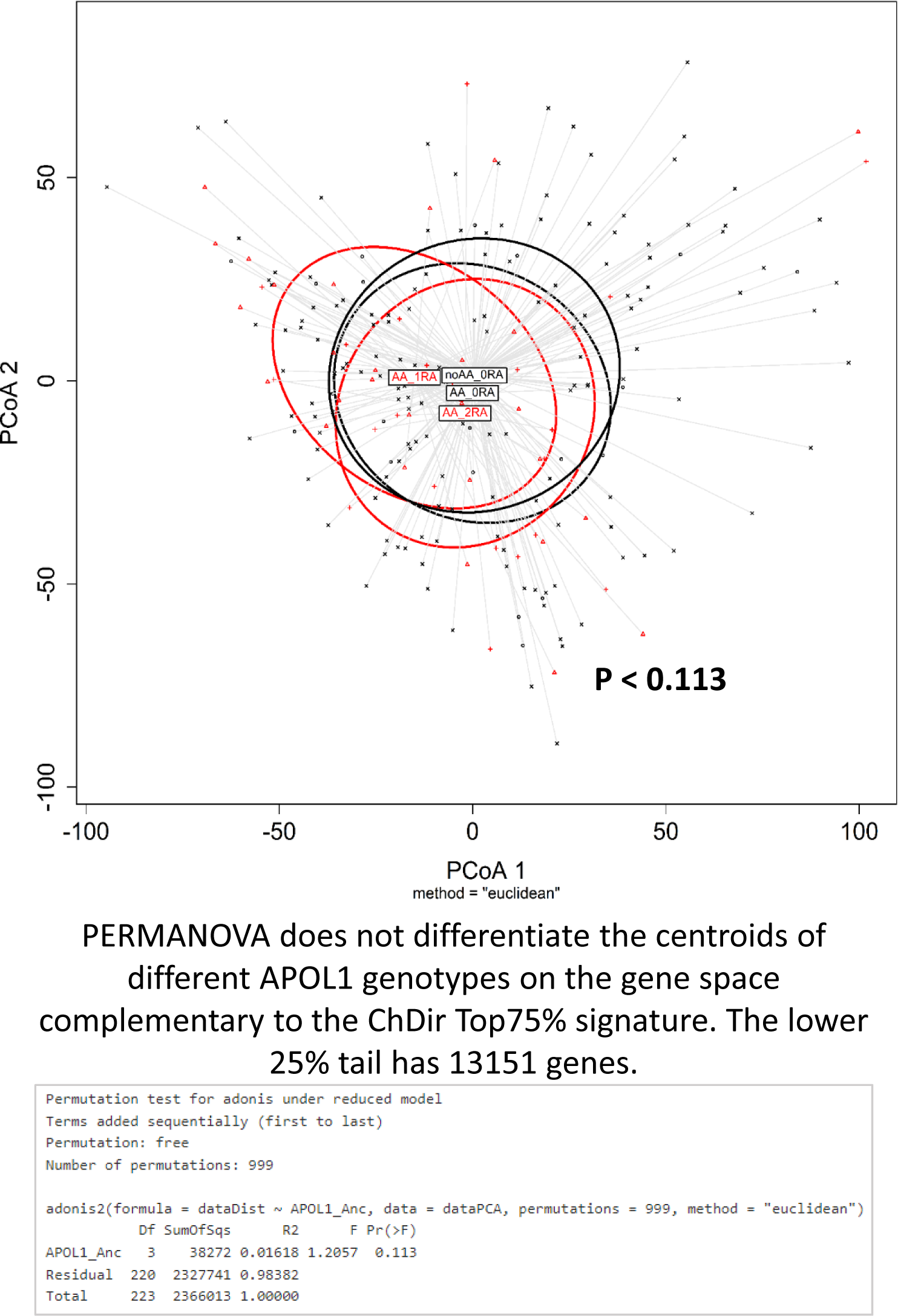
Principal Coordinate Analysis using the Euclidean distance matrices of the genes complementary to ChDir. PERMANOVA failed to separate NEPTUNE participants by *APOL1* risk allele number within this gene space (13,151 genes).

**Supplemental Figure 6:**
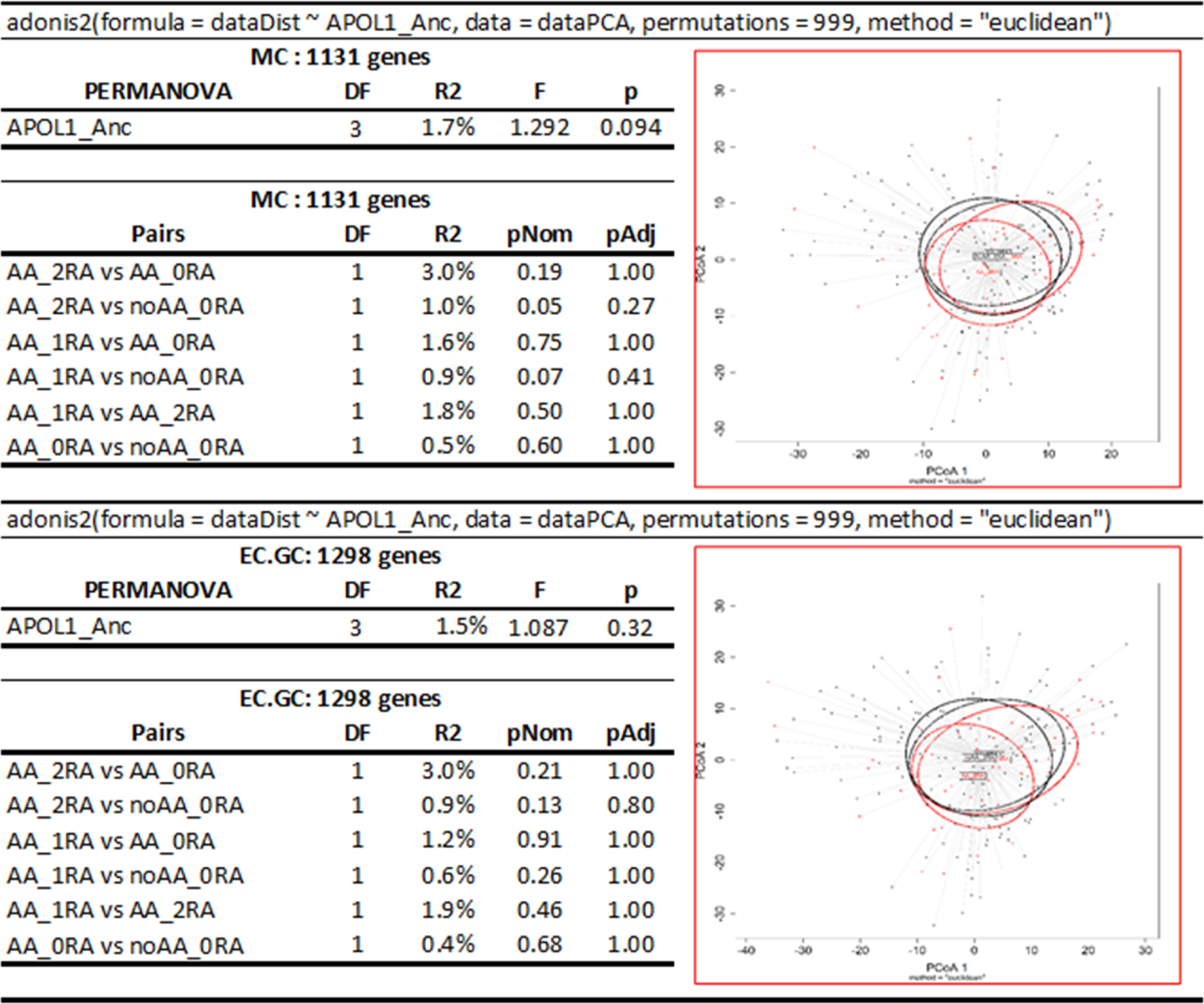
PERMANOVA of NEPTUNE participants using the cell identity signatures for mesangial cells (MC) and glomerular endothelial cells (EC.GC gene). PERMANOVA and Principal Coordinates Analysis using the Euclidean distance matrices of cell-identity signatures for the glomerular mesangium (MC) and the glomerular capillary endothelium (EC-GC) show lack of separation between NEPTUNE participants with different *APOL1* genotypes.

## SUPPLEMENTAL TABLES

**Table S1:** *APOL1* alleles in AA stratified by kidney biopsy histopathology.

| **Diagnosis**<br>**(Cohort)** | **APOL1 risk alleles** |  |  | **Total**<br>**(#)** |
| --- | --- | --- | --- | --- |
|  | 2RA | 1RA | 0RA |  |
| **FSGS** | 17 | 14 | 4 | 35 |
| **MCD** | 4 | 9 | 11 | 24 |
| **MN** | 1 | 7 | 5 | 13 |
| **Total (#)** | 22 | 30 | 20 | 72 |
| **Total (%)** | 30.6% | 41.7% | 27.8% |  |

**Table S2:** WGCNA Network modules and metamodules gene assignments (Provided as *.csv file)

**Table S3:** Characteristic direction gene signature and ranks for AA_2RA vs AA_0RA (Provided as *.xlsx file)

**Table S4:** Kidney Precision Medicine Project single-cell and single-nuclei RNAseq data integration (Provided as *.xlsx file)

**Table S5:** Number of genes in the Cell-Identity and Cell-Exclusive gene signatures.

| Anatomy as defined by the KPMP |  |  | Cell-identity<br>gene signature | Cell-exclusive<br>gene signature |
| --- | --- | --- | --- | --- |
| Region | subRegion | subRegion.ID | (#) | (#) |
| Glomerulus / Renal Corpuscle | Glomerular Visceral Epithelium | POD | 1814 | 125 |
| Glomerulus / Renal Corpuscle | Glomerular Parietal Epithelium | PEC | 1220 | 26 |
| Glomerulus / Renal Corpuscle | Glomerular Mesangium | MC | 1131 | 15 |
| Glomerulus / Renal Corpuscle | Glomerular Capillary Endothelium | ECGC | 1298 | 9 |
| Tubules | Proximal Tubule | PT | - | 425 |
| Tubules | Loop of Henle (Thin Limb) | Thin.Limbs | - | 201 |
| Tubules | Loop of Henle (Thick Limb) | TAL | - | 80 |
| Tubules | Distal Convolution | DCT | - | 131 |
| Tubules | Connecting Tubule | CNT | - | 88 |
| Tubules | Collecting Duct | CD | - | 132 |
| Vessels | Endothelium (Non-glomerular) | EC | - | 165 |
| Interstitium | Stroma (Non-glomerular) | SC | - | 365 |
| Interstitium | Immune | IMN | - | 950 |

## EXTENDED METHODS

### NEPTUNE Study Cohort

The Nephrotic Syndrome Study Network (NEPTUNE) is a longitudinal study of patients with primary nephrotic syndrome [1]. Participants in the NEPTUNE Biopsy Cohort provide a biopsy core at enrollment, as well as blood, urine, and clinical data at enrollment and at 4 to 6 month intervals. The NEPTUNE Data Coordination Center controls access to the data and provided RNAseq glomerular batch-corrected count matrices from 224 individuals with associated clinical data, including *APOL1* genotypes and clinical outcomes. Data is available from NEPTUNE upon request.

A variable called inferred African ancestry (AA) was defined positive for those patients having an *APOL1* genotype different from G0/G0, and/or a self-reported race of “Black/African American” (n=72); while G0/G0 individuals, who did not self-identify as “Black/African American,” were defined as not having African ancestry (noAA, n=152). Self-reported race and presence of *APOL1* risk alleles highly correlated with African genetic ancestry in NEPTUNE participants [2, 3].

### NEPTUNE glomerular RNAseq data

Glomerular transcriptomes from microdissected glomeruli were obtained from the NEPTUNE consortium with their associated clinical data. All available glomerular transcriptomes (n=224) from the NEPTUNE study cohort were quality controlled and normalized as a group yielding 14632 protein-coding genes. A geneINFO data frame was obtained using the R package Annotables and the grch38 genome information. The R package DESeq2 was used for quality control and normalization of count tables. Genes with 0 (zero) variance were eliminated and a DESeq2 object was created. Sequencing depth was normalized using the function *estimateSizeFactors()*, and size factors were saved to the Clinical/metadata data frame. Principal Component Analysis (PCA) was conducted using R packages FactoMineR and factoextra for quality control. Two sample outliers were identified and excluded from the remaining analyses, after which the raw data was renormalized and re-inspected. Protein-coding genes with valid NCBI IDs were selected from the normalized count matrix, which yielded 14632 genes. One count was added to all values on the matrix and then a Log_2_ transformation was applied. Data was re-inspected using boxplots and PCA. The Normalized/Filtered/Log_2_-transformed matrix, and the relational Clinical Data and geneINFO data frames were saved into an R object for downstream analyses. In some analyses, the Normalized/Filtered/Log_2_-transformed matrix was adjusted for estimated glomerular filtration rate (eGFR) and urinary protein creatinine ratio (UPCR) (∼ eGFR + UPCR), by fitting a linear model in the R package Limma [4] and extracting the residuals matrix.

### *APOL1* transgenic mouse glomerular microarray data

We generated glomerular transcriptomes from 40-day old Tg26 transgenic mice with *APOL1*-G0 or *APOL1*-G2 transgenes controlled by the murine *Nphs1* promoter [5, 6]. Glomeruli were isolated by successive sieving as previously described [7]. Total RNA was extracted from the glomeruli using methods similar than before [8–10]. In brief, total RNA was extracted using RNeasy mini kit (QIAGEN) according to manufacturer’s recommendations, quantitated using a NanoDrop 2000 spectrophotometer (Thermo Scientific), and adjusted to a final concentration of 50 ng/μl with nuclease free water (Qiagen). The concentration-adjusted RNA was submitted to the Genomics Core at CWRU School of Medicine. RNA expression data were generated using the Mouse Gene 2.0 ST Array (Affymetrix). Individual cluster transcript IDs were mapped to the mouse genome, summarized and annotated using 23748 mouse EntrezIDs. Batch correction was performed using ComBat [11]. Genes that were not expressed above background in at least 50% of the samples were removed, yielding a total of 19170 annotated genes. The R package Limma [4] was used to fit a linear model for sex (∼sex), and the residuals matrix was extracted for analysis.

### Weighted Gene Co-Expression Network Analysis (WGCNA) in individuals with AA

The 72 individuals with AA were selected to build a gene coexpression network. PCA did not identify any trait in the clinical data set that clustered the genomewide transcriptomes. A gene coexpression network was constructed using WGCNA methodology, as implemented in the R package WGCNA [12, 13]. WGCNA consists of two main steps. In the first step, a coexpression network is constructed from gene expression data. For this, a Pearson-correlation coefficient is calculated for each gene pair using the gene expression pair values from each sample as individual points. The correlation coefficient of all samples is then stored in a symmetric matrix (#genes, #genes). The correlation matrix is transformed into an adjacency matrix by applying the adjacency function, and then into the topological overlap matrix (TOM) by counting the n-step connections shared by any pair of genes [14]. We used the “signed” version of the adjacency function and selected the soft thresholding power accordingly to the scale-free topology criterion [14]. Finally, genes with shared connectivity were clustered into coexpression modules. WGCNA summarizes the expression profiles of the genes within each coexpression module (in-module genes) into one vector called a module eigengene, which weights the expression of all genes within a module for each sample. The second step in WGCNA calculates if the module eigengene correlates with a trait of interest by using the eigengene/trait pairs for each sample. For instance, to explore the significance of a module with FSGS, a discrete variable corresponding to a histopathological classification was created. Patients were coded as “1” if they were diagnosed with FSGS or as “0” if they were diagnosed with either MCD or MN. Then the module significance was calculated as the Pearson correlation coefficient between the eigengene of each sample and FSGS [13, 15]. The significance threshold was adjusted by the number of coexpression modules using the Bonferroni method. This procedure can also be implemented with continuous variables such as eGFR.

### Correlation of transcriptomics analyses with clinical outcomes

NEPTUNE participants are followed prospectively for up to five years, providing longitudinal data for the following clinical outcomes: 1) “Complete Remission”, defined as UPCR≤0.3 at any visit after screening among patients with active disease at screening; and 2) “Composite Event” defined as a patient reaching ESKD (two consecutive eGFR < 15, dialysis or transplantation) or an eGFR <90 and a decline ≥40% from baseline. We conducted Kaplan-Meier analysis using gene expression z-scores tertiles as surrogates for the gene activation signatures of interest at the time of kidney biopsy as previously reported [16, 17]. In brief, gene expression levels were z-transformed; then, an average z-score for each gene signature was calculated for the 191 NEPTUNE participants (60 AA and 161 noAA) with sufficient longitudinal data to assess outcomes. Finally, patients were stratified by gene activation tertiles [high, medium, low]. Differences between the curves were tested using the log-rank test. We conducted Kaplan-Meier analysis using the following gene spaces: 1) MM2 (437 genes), 2) Midnightblue module (226 genes) and 3) the *APOL1* ChDir signature (1481 genes). Independent analyses were conducted on AA and nonAA NEPTUNE participants.

### Functional enrichment analyses

We utilized EnrichR [18, 19] to conduct functional enrichment analysis using the Molecular Signatures Database (MSigDB) Hallmark gene sets [20]. The Hallmark gene sets “summarize and represent specific well-defined biological states or processes and display coherent expression. These gene sets were generated by a computational methodology based on identifying overlaps between gene sets in other MSigDB collections and retaining genes that display coordinate expression” [21].

### Characteristic Direction (ChDir)

ChDir [22] is a geometric multivariate approach to differential gene expression and was previously used by us and other investigators to obtain transcriptional signatures from proximal tubules [10, 23], podocytes [24], and human kidney cancerous cells [25]. This method generates a hyperplane that separates two classes in an n_(gene)_-dimensional space. The hyperplane orientation, defined by a normal vector, describes overall differences in gene expression between two conditions. Variance shrinkage gives more weight to coexpressed genes than to those with large expression differences between classes. Expression differences are normalized, so that the sum of the squares of all genes adds up to 1 and then ranked in descending order. These normalized gene vectors represent the fractional contribution of each gene to the overall transcriptional differences between classes, which allows the extraction of the top-scoring classifier genes accounting for 75% of discrimination between classes. In our analyses, these genes represent a signature that differentiates individual by number of *APOL1* kidney risk alleles.

### Single-cell (sc) and single-nucleus (sn) RNAseq matrices

We previously used the Kidney Precision Medicine Project (KPMP) scRNAseq and snRNAseq data [26–28] to obtain quantitative full transcriptomes [29] as well as gene enrichment [9] of kidney cells. In these approaches, some genes are enriched in several cell types and can reduce the accuracy of cell identity signatures. To mitigate this problem, we used a stringent method to obtain cell identity signatures enriched in specific glomerular cells, as well as cell identity signatures unique to specific kidney cell types.

We downloaded single cell (scRNAseq) and single nuclear (snRNAseq) RNA sequencing data from the Kidney Tissue Atlas (https://atlas.kpmp.org/explorer/) during September and October 2021. The scRNAseq expression dataset included 12 AKI and 15 CKD participants, and the snRNAseq expression dataset included 3 healthy reference tissue, 6 AKI and 10 CKD participants. Together, both technologies detected 85 non-redundant clusters from 13 anatomical sub-regions defined by the KPMP as: 1) Glomerular Visceral Epithelium (POD), 2) Glomerular Parietal Epithelium (PEC), 3) Glomerular Mesangium (MC), 4) Glomerular Capillary Endothelium (ECGC), 5) Proximal Tubule (PT), 6) Loop of Henle thin portion (Thin Limbs), 7) Loop of Henle thick portion (TAL), 8) Distal Convolution (DCT), 9) Connecting Tubule (CNT), 10) Collecting Duct (CD), 11) Endothelium Non-Glomerular (EC), 12) Stroma Non-Glomerular (SC), and 13) Immune (IMN) (**Supplemental Table 3; Tab: Metadata**).

The data integration workflow can be found in **Supplemental Table 3; Tab: Data Integration** and Followed multiple steps: <u>Step 1</u>) a matrix with dimensions (i x j), corresponding to genes (i) and cell-clusters (j) was created for each technology, i.e. scRNAseq and snRNAseq. Each of these matrices contained either a positive log_2_ fold-change (FC) value for the genes expressed in each cluster compared to the average of all other clusters, which are defined as non-enriched genes (NS); <u>Step 2</u>) both matrices were expanded to match the gene and cluster dimensions by imputing ‘NA’ in all new fields. Then, matrices were transformed to binary expression matrices by assigning a value of ‘1’ for any value larger than zero (>0) and a value of ‘0’ for “NS” and “NA” cells; <u>Step 3)</u> A “Binary Sum” matrix was created by the element wise addition of both binary matrices. The possible values of the binary sum matrix were ‘0’, ‘1’ or ‘2’, indicating that the expression of element ‘i’ (gene) was found above average in the ‘j’ (cell-cluster) by ‘none’, ‘one’ or ‘two’ sequencing technologies, respectively. <u>Step 4</u>) a subRegion expression signature matrix was created by grouping the expression of individual cell clusters by anatomical sub-regions as defined above. As an example, the sub-region Glomerular Visceral Epithelium (POD) integrates the data from both technologies, and contains all genes with a positive fold change in either podocytes or degenerative podocytes. Finally, the 14617 genes that mapped to the NEPTUNE network genes (14632 genes) were annotated and selected for downstream analysis (**Supplemental Table 3; Tab: subRegion expression matrix**). The total number of genes on each of the cell identity and cell exclusive gene signatures can be found in **Supplemental Table 3; Tab: Cell Signatures**.

### Permutational Multivariate Analysis of Variance (PERMANOVA) and Principal Coordinates Analysis (PCoA)

We used PERMANOVA [30, 31] to determine the statistical significance of differences between two or more groups of transcriptomes based on gene expression data. PERMANOVA evaluates group separations by calculating a pseudo-F statistic, comparing the variability within groups to the variability between groups, based on a specified distance matrix. The method provides an R² statistic, representing the percentage of variability explained by group differences, and a p-value, indicating the statistical significance of the separation. To implement this method, we applied the function *adonis2()* from the R package ‘vegan’ [32]. We inputted the Euclidian distance matrix as it emphasizes the actual proximity of gene expression values [33] as opposed to WGCNA and ChDir, which are leveraged towards covariance analysis. This approach allowed us to look at different aspects of the transcriptional landscape. In cases where more than two groups were compared, we performed post-hoc pairwise comparisons using the *pairwise.adonis()* function with default parameters [81] to adjust the p-values. Other investigators used PERMANOVA for transcriptome analysis before [36, 37], including in kidney transcriptomics [34, 35]. To visualize the separation of groups, we performed Principal Coordinates Analysis (PCoA) using the Euclidean distance matrix. PCoA reduces the dimensionality of the data, allowing us to represent the relationships between transcriptomes graphically while preserving as much distance information as possible. The first two principal coordinates were used to generate the ordination plots shown in the figures.

**Module Scores:** gene module expression scores in kidney cells were calculated as previously reported [8]. In brief, the “Integrated Single-nucleus and Single-cell RNA-seq of the Adult Human Kidney” (GSE169285) dataset was downloaded on December 8^th^ 2023, from https://cellxgene.cziscience.com/collections/bcb61471-2a44-4d00-a0af-ff085512674c. The downloaded file name was “baa97c56-c7a0-4858-bdcd-3fb5802177ed.rds”, which contains transcriptomes from 304,652 cells from the kidney cortex, medulla, and papilla. Analyses were conducted using the R package Seurat (version 5.1.0). Glomerular cells were extracted based on the subclass.l2 annotation, yielding 3,764 EC-GC, 458 MC, 2,420 POD, 412 degenerative POD (dPOD), and 2,417 PEC. Genes associated with MM2, MM4, and MM10 were mapped onto glomerular cell clusters, and module scores were calculated using the *AddModuleScore()* function and visualized using *VlnPlot()*.

## Notes

### Competing Interest Statement

The authors have declared no competing interest.

### Author Declarations

The NEPTUNE protocol is approved by the University of Michigan IRBMED central IRB (HUM00158219), and the Cleveland Clinic IRB (15-182). The KPMP study protocol is approved using a central IRB at the Human Research Protection Office of Washington University in St. Louis (IRB no. 201902013). NEPTUNE and KPMP participants provided written informed consent prior to enrollment.

